# Burden of lower respiratory tract infections preventable by adult immunization with 15- and 20-valent pneumococcal conjugate vaccines in the United States

**DOI:** 10.1101/2023.02.23.23286380

**Authors:** Joseph A. Lewnard, Vennis Hong, Katia J. Bruxvoort, Lindsay R. Grant, Luis Jódar, Alejandro Cané, Adriano Arguedas, Magdalena E. Pomichowski, Bradford D. Gessner, Sara Y. Tartof

## Abstract

**Background:** Updated 2022 recommendations indicate all US adults aged ≥65 years and adults aged <65 years with comorbid conditions should receive 15- and 20-valent pneumococcal conjugate vaccines (PCV15/20). We aimed to assess the potential impact of these recommendations on the burden of lower respiratory tract infections (LRTIs) among adults.

**Methods:** We estimated the incidence of LRTI cases and associated hospital admissions among enrollees of Kaiser Permanente Southern California health plans from 2016-19. We used a counterfactual inference framework to estimate excess LRTI-associated risk of death up to 180 days after diagnosis. We used prior estimates of PCV13 effectiveness against all-cause and serotype-specific LRTI to model potential direct effects of PCV15/20 by age group and risk status.

**Results:** Use of PCV15 and PCV20, respectively, could prevent 89.3 (95% confidence interval: 41.3-131.8) and 108.6 (50.4-159.1) medically-attended LRTI cases per 10,000 person-years; 21.9 (10.1-32.0) and 26.6 (12.4-38.7) hospitalized LRTI cases per 10,000 person-years; and 7.1 (3.3-10.5) and 8.7 (4.0-12.7) excess LRTI-associated deaths per 10,000 person-years. Among at-risk adults aged <65 years not previously prioritized for receipt of PCV13, PCV15 and PCV20, respectively, could prevent 85.7 (39.6-131.5) and 102.7 (47.8-156.7) medically-attended LRTI cases per 10,000 person-years; 5.1 (2.4-8.6) and 6.2 (2.8-10.2) LRTI hospitalizations per 10,000 person-years, and 0.9 (0.4-1.4) and 1.1 (0.5-1.7) excess LRTI-associated deaths per 10,000 person-years. Expansions in serotype coverage, relative to PCV13, accounted for the majority of the expected increase in vaccine-preventable hospitalizations and deaths.

**Conclusions:** Our findings suggest recent recommendations including PCV15/20 within adult pneumococcal vaccine series may substantially reduce LRTI burden.

**Key points:**

- Use of PCV15/20 among US adults aged ≥65 years may prevent 521,000-626,000 LRTI cases, 127,000-154,000 hospitalizations, and 50,000-61,000 excess deaths, annually.
- Updated recommendations for PCV15/20 among adults aged <65 years may prevent 441,000-526,000 LRTI cases, 39,000-46,000 hospitalizations, and 8,000-9,000 deaths annually.

## INTRODUCTION

Lower respiratory tract infections (LRTIs) including community-acquired pneumonia (CAP) are a leading cause of severe illness, healthcare utilization, and mortality among adults.^1^ *Streptococcus pneumoniae* (pneumococcus) is a prominent LRTI etiology associated with increased likelihood of CAP presentation and fatal disease outcomes.^2^ Since 1997, all US adults aged ≥65 years have been recommended to receive pneumococcal polysaccharide vaccine (PPSV23) containing capsular antigens from 23 pneumococcal serotypes to prevent invasive pneumococcal disease (IPD), including bacteremic pneumonia, caused by vaccine-targeted serotypes.^3^ However, PPSV23 has not consistently been found to protect against non-invasive infections, which account for most pneumococcal LRTI cases. In 2014, a randomized controlled trial in the Netherlands reported that 13-valent pneumococcal conjugate vaccine (PCV13) prevents nonbacteremic CAP caused by vaccine-targeted pneumococcal serotypes among adults aged ≥65 years.^4^ Postlicensure studies have confirmed PCV13 effectiveness against non-bacteremic CAP due to vaccine-targeted pneumococcal serotypes among US older adults,^5^ and estimated that direct vaccination with PCV13 reduces risk of all-cause LRTI or CAP by 7-12% among older adults in settings with established pediatric PCV13 programs.^6–10^

The US Advisory Committee on Immunization Practices (ACIP) recommended in 2014 that all adults aged ≥65 years, and high-risk adults aged <65 years with immunocompromising conditions, functional or anatomical asplenia, cerebrospinal fluid leak, or cochlear implants, should receive PCV13 (targeting serotypes 1, 3, 4, 5, 6A, 6B, 7F, 9V, 14, 18C, 19A, 19F, 23F) in succession with PPSV23 to prevent CAP and IPD.^11^ In 2022, newer-generation PCVs targeting 15 and 20 pneumococcal serotypes (PCV15/20; adding 22F and 33F [both vaccines] and 8, 10A, 11A, 12F, and 15B [PCV20 only]) were licensed for use among older adults.^12,13^ Updated ACIP recommendations in 2022 stated all PCV-naïve adults aged ≥65 years should receive PCV15 in sequence with PPSV23, or PCV20 alone, to prevent CAP and IPD.^14^ This update also expanded PCV15+PPSV23 or PCV20 recommendations to a broad population of “at-risk” adults aged <65 years previously recommended to receive PPSV23 without PCV13, including those with chronic heart, liver, or lung diseases; diabetes; and a history of alcoholism or cigarette smoking.

Whereas US sentinel surveillance programs monitor IPD incidence and serotype distribution,^15^ the burden of non-bacteremic CAP and other LRTIs preventable by PCV15/20 remains unknown. Using data from a large healthcare system in California, we aimed to estimate potential impacts of transitioning from PCV13 to PCV15/20, and the expansion of PCV recommendations to at-risk adults aged <65 years, on the incidence of medically-attended LRTI, associated hospital admissions, and mortality.

## METHODS

### Setting

The Kaiser Permanente Southern California (KPSC) healthcare system serves ∼4.7 million members (∼19% of the population of greater Los Angeles, San Diego, and surrounding areas), broadly representing the region’s racially and ethnically diverse population.^16^ Plans include employer-provided, pre-paid, or federally-sponsored health insurance coverage mechanisms. Healthcare delivery across virtual, outpatient, emergency department (ED), and inpatient settings, including diagnoses, provider notes, laboratory tests and results, and prescriptions, is captured by patient electronic health records (EHRs). Out-of-network care is tracked through insurance claims, enabling near-complete capture of healthcare interactions.

### Design

Eligible individuals were aged ≥18 years and belonged to KPSC health plans between 1 January, 2016 and 31 December, 2019. Data from 2020 onward were excluded due to the COVID-19 pandemic. We monitored outcomes within the study population during each calendar year, censoring individuals at death or disenrollment. Individuals contributed observations in calendar years preceded by ≥1 year of continuous coverage (allowing for membership lapses of ≤45 days). The primary endpoint was new-onset LRTI (without a previous diagnosis within 30 days) due to any cause and diagnosed in any setting (**Table S1**). We distinguished LRTI cases diagnosed with CAP or other LRTI diagnoses, and cases diagnosed in inpatient or outpatient (including ED) settings. Cases with multiple CAP or non-CAP LRTI diagnoses <30 days apart were considered to have experienced a single LRTI episode.

### Incidence

We computed incidence rates for all outcomes stratified by ACIP-designated risk group^17^ and age (18-29, 30-49, 50-64, 65-79, and ≥80 years). Risk groups included: (1) “low-risk” immunocompetent individuals without major comorbid conditions; (2) “at-risk” immunocompetent individuals with comorbid conditions associated with increased likelihood of pneumococcal disease (alcoholism; chronic heart diseases including congestive heart failure and cardiomyopathies, but excluding hypertension; chronic lung diseases including chronic obstructive pulmonary disease, emphysema, and asthma; diabetes mellitus; chronic liver disease or cirrhosis; and cigarette smoking); and (3) “high-risk” individuals with cerebrospinal fluid leak, cochlear implant, functional or anatomic asplenia, or any immunocompromising condition (including congenital or acquired immunodeficiencies, HIV infection, chronic renal failure, nephrotic syndrome, leukemia, lymphoma, Hodgkin disease, malignancy, solid organ transplant, multiple myeloma, or any iatrogenic immunosuppression, including via systemic corticosteroids or radiation therapy). We estimated rates within each stratum by fitting Poisson regression models to case counts at the person-year level, defining cluster-robust standard errors to accommodate multiple observation-years per individual. Models included calendar-year intercepts. We sampled from the fitted distribution of rate parameters across all years to propagate statistical uncertainty and year-to-year variability in incidence projections.

### Excess mortality

We used a counterfactual inference framework to quantify excess LRTI-associated mortality over 30, 60, 90, and 180-day intervals among new-onset CAP and non-CAP LRTI cases. We describe the statistical approach in the supporting information (**Text S1**). Briefly, we compared observed mortality among cases to same-season mortality among matched individuals without contemporaneous LRTI diagnoses. We matched cases and non-LRTI controls on age group, sex, race/ethnicity, body mass index, cigarette smoking, neighborhood socioeconomic status, ACIP-defined risk group, history of healthcare utilization across outpatient, inpatient, and ED settings in the prior year, history of LRTI in the prior year, vaccines received, and comorbid conditions (as categorized in **Table 1**).

**Table 1:**
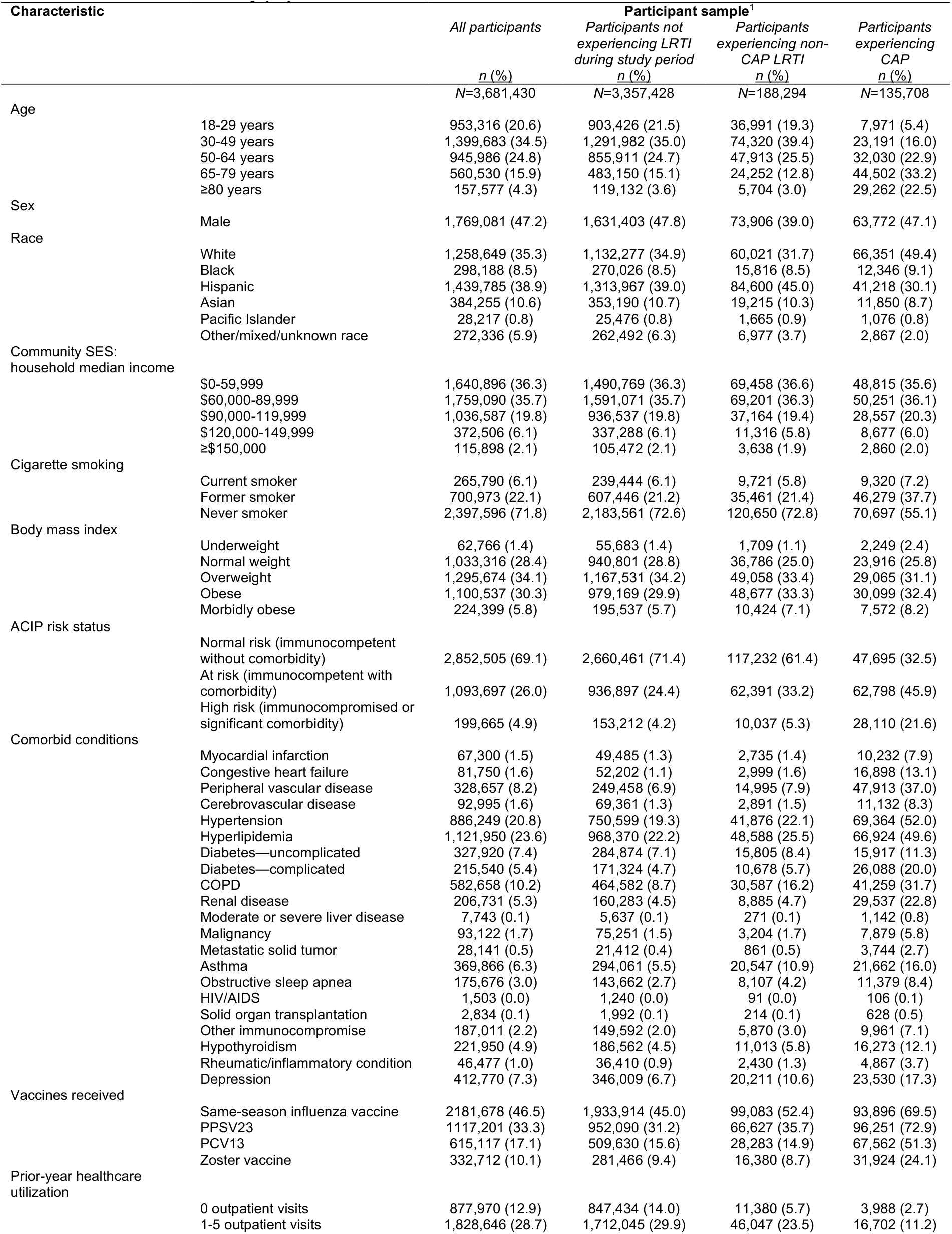
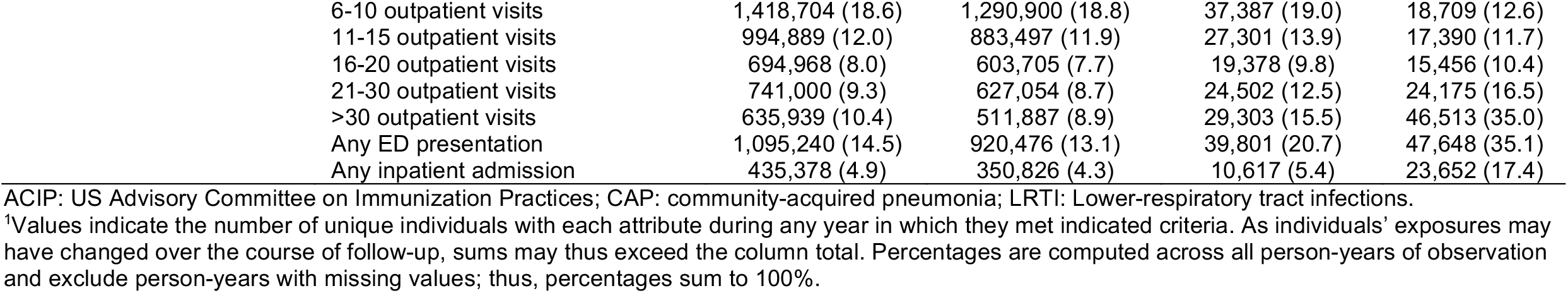
Attributes of the study population.

| Characteristic |  | Participant sample<sup>1</sup> |  |  |  |
| --- | --- | --- | --- | --- | --- |
|  |  | All participants | Participants not experiencing LRTI during study period | Participants experiencing non-CAP LRTI | Participants experiencing CAP |
|  |  | *n* (%) | *n* (%) | *n* (%) | *n* (%) |
|  |  | N=3,681,430 | N=3,357,428 | N=188,294 | N=135,708 |
| Age | 18-29 years | 953,316 (20.6) | 903,426 (21.5) | 36,991 (19.3) | 7,971 (5.4) |
|  | 30-49 years | 1,399,683 (34.5) | 1,291,982 (35.0) | 74,320 (39.4) | 23,191 (16.0) |
|  | 50-64 years | 945,986 (24.8) | 855,911 (24.7) | 47,913 (25.5) | 32,030 (22.9) |
|  | 65-79 years | 560,530 (15.9) | 483,150 (15.1) | 24,252 (12.8) | 44,502 (33.2) |
|  | ≥80 years | 157,577 (4.3) | 119,132 (3.6) | 5,704 (3.0) | 29,262 (22.5) |
| Sex | Male | 1,769,081 (47.2) | 1,631,403 (47.8) | 73,906 (39.0) | 63,772 (47.1) |
| Race | White | 1,258,649 (35.3) | 1,132,277 (34.9) | 60,021 (31.7) | 66,351 (49.4) |
|  | Black | 298,188 (8.5) | 270,026 (8.5) | 15,816 (8.5) | 12,346 (9.1) |
|  | Hispanic | 1,439,785 (38.9) | 1,313,967 (39.0) | 84,600 (45.0) | 41,218 (30.1) |
|  | Asian | 384,255 (10.6) | 353,190 (10.7) | 19,215 (10.3) | 11,850 (8.7) |
|  | Pacific Islander | 28,217 (0.8) | 25,476 (0.8) | 1,665 (0.9) | 1,076 (0.8) |
|  | Other/mixed/unknown race | 272,336 (5.9) | 262,492 (6.3) | 6,977 (3.7) | 2,867 (2.0) |
| Community SES: household median income | \$0-59,999 | 1,640,896 (36.3) | 1,490,769 (36.3) | 69,458 (36.6) | 48,815 (35.6) |
| | \$60,000-89,999 | 1,759,090 (35.7) | 1,591,071 (35.7) | 69,201 (36.3) | 50,251 (36.1) |
| | \$90,000-119,999 | 1,036,587 (19.8) | 936,537 (19.8) | 37,164 (19.4) | 28,557 (20.3) |
| | \$120,000-149,999 | 372,506 (6.1) | 337,288 (6.1) | 11,316 (5.8) | 8,677 (6.0) |
| | ≥\$150,000 | 115,898 (2.1) | 105,472 (2.1) | 3,638 (1.9) | 2,860 (2.0) |
| Cigarette smoking | Current smoker | 265,790 (6.1) | 239,444 (6.1) | 9,721 (5.8) | 9,320 (7.2) |
|  | Former smoker | 700,973 (22.1) | 607,446 (21.2) | 35,461 (21.4) | 46,279 (37.7) |
|  | Never smoker | 2,397,596 (71.8) | 2,183,561 (72.6) | 120,650 (72.8) | 70,697 (55.1) |
| Body mass index | Underweight | 62,766 (1.4) | 55,683 (1.4) | 1,709 (1.1) | 2,249 (2.4) |
|  | Normal weight | 1,033,316 (28.4) | 940,801 (28.8) | 36,786 (25.0) | 23,916 (25.8) |
|  | Overweight | 1,295,674 (34.1) | 1,167,531 (34.2) | 49,058 (33.4) | 29,065 (31.1) |
|  | Obese | 1,100,537 (30.3) | 979,169 (29.9) | 48,677 (33.3) | 30,099 (32.4) |
|  | Morbidly obese | 224,399 (5.8) | 195,537 (5.7) | 10,424 (7.1) | 7,572 (8.2) |
| ACIP risk status | Normal risk (immunocompetent without comorbidity) | 2,852,505 (69.1) | 2,660,461 (71.4) | 117,232 (61.4) | 47,695 (32.5) |
|  | At risk (immunocompetent with comorbidity) | 1,093,697 (26.0) | 936,897 (24.4) | 62,391 (33.2) | 62,798 (45.9) |
|  | High risk (immunocompromised or significant comorbidity) | 199,665 (4.9) | 153,212 (4.2) | 10,037 (5.3) | 28,110 (21.6) |
| Comorbid conditions | Myocardial infarction | 67,300 (1.5) | 49,485 (1.3) | 2,735 (1.4) | 10,232 (7.9) |
|  | Congestive heart failure | 81,750 (1.6) | 52,202 (1.1) | 2,999 (1.6) | 16,898 (13.1) |
|  | Peripheral vascular disease | 328,657 (8.2) | 249,458 (6.9) | 14,995 (7.9) | 47,913 (37.0) |
|  | Cerebrovascular disease | 92,995 (1.6) | 69,361 (1.3) | 2,891 (1.5) | 11,132 (8.3) |
|  | Hypertension | 886,249 (20.8) | 750,599 (19.3) | 41,876 (22.1) | 69,364 (52.0) |
|  | Hyperlipidemia | 1,121,950 (23.6) | 968,370 (22.2) | 48,588 (25.5) | 66,924 (49.6) |
|  | Diabetes—uncomplicated | 327,920 (7.4) | 284,874 (7.1) | 15,805 (8.4) | 15,917 (11.3) |
|  | Diabetes—complicated | 215,540 (5.4) | 171,324 (4.7) | 10,678 (5.7) | 26,088 (20.0) |
|  | COPD | 582,658 (10.2) | 464,582 (8.7) | 30,587 (16.2) | 41,259 (31.7) |
|  | Renal disease | 206,731 (5.3) | 160,283 (4.5) | 8,885 (4.7) | 29,537 (22.8) |
|  | Moderate or severe liver disease | 7,743 (0.1) | 5,637 (0.1) | 271 (0.1) | 1,142 (0.8) |
|  | Malignancy | 93,122 (1.7) | 75,251 (1.5) | 3,204 (1.7) | 7,879 (5.8) |
|  | Metastatic solid tumor | 28,141 (0.5) | 21,412 (0.4) | 861 (0.5) | 3,744 (2.7) |
|  | Asthma | 369,866 (6.3) | 294,061 (5.5) | 20,547 (10.9) | 21,662 (16.0) |
|  | Obstructive sleep apnea | 175,676 (3.0) | 143,662 (2.7) | 8,107 (4.2) | 11,379 (8.4) |
|  | HIV/AIDS | 1,503 (0.0) | 1,240 (0.0) | 91 (0.0) | 106 (0.1) |
|  | Solid organ transplantation | 2,834 (0.1) | 1,992 (0.1) | 214 (0.1) | 628 (0.5) |
|  | Other immunocompromise | 187,011 (2.2) | 149,592 (2.0) | 5,870 (3.0) | 9,961 (7.1) |
|  | Hypothyroidism | 221,950 (4.9) | 186,562 (4.5) | 11,013 (5.8) | 16,273 (12.1) |
|  | Rheumatic/inflammatory condition | 46,477 (1.0) | 36,410 (0.9) | 2,430 (1.3) | 4,867 (3.7) |
|  | Depression | 412,770 (7.3) | 346,009 (6.7) | 20,211 (10.6) | 23,530 (17.3) |
| Vaccines received | Same-season influenza vaccine | 2181,678 (46.5) | 1,933,914 (45.0) | 99,083 (52.4) | 93,896 (69.5) |
|  | PPSV23 | 1117,201 (33.3) | 952,090 (31.2) | 66,627 (35.7) | 96,251 (72.9) |
|  | PCV13 | 615,117 (17.1) | 509,630 (15.6) | 28,283 (14.9) | 67,562 (51.3) |
|  | Zoster vaccine | 332,712 (10.1) | 281,466 (9.4) | 16,380 (8.7) | 31,924 (24.1) |
| Prior-year healthcare utilization | 0 outpatient visits | 877,970 (12.9) | 847,434 (14.0) | 11,380 (5.7) | 3,988 (2.7) |
|  | 1-5 outpatient visits | 1,828,646 (28.7) | 1,712,045 (29.9) | 46,047 (23.5) | 16,702 (11.2) |

|  |  |  |  |  |
| --- | --- | --- | --- | --- |
| 6-10 outpatient visits | 1,418,704 (18.6) | 1,290,900 (18.8) | 37,387 (19.0) | 18,709 (12.6) |
| 11-15 outpatient visits | 994,889 (12.0) | 883,497 (11.9) | 27,301 (13.9) | 17,390 (11.7) |
| 16-20 outpatient visits | 694,968 (8.0) | 603,705 (7.7) | 19,378 (9.8) | 15,456 (10.4) |
| 21-30 outpatient visits | 741,000 (9.3) | 627,054 (8.7) | 24,502 (12.5) | 24,175 (16.5) |
| >30 outpatient visits | 635,939 (10.4) | 511,887 (8.9) | 29,303 (15.5) | 46,513 (35.0) |
| Any ED presentation | 1,095,240 (14.5) | 920,476 (13.1) | 39,801 (20.7) | 47,648 (35.1) |
| Any inpatient admission | 435,378 (4.9) | 350,826 (4.3) | 10,617 (5.4) | 23,652 (17.4) |

ACIP: US Advisory Committee on Immunization Practices; CAP: community-acquired pneumonia; LRTI: Lower-respiratory tract infections.
<sup>1</sup>Values indicate the number of unique individuals with each attribute during any year in which they met indicated criteria. As individuals' exposures may have changed over the course of follow-up, sums may thus exceed the column total. Percentages are computed across all person-years of observation and exclude person-years with missing values; thus, percentages sum to 100%.

### Vaccine impact

We estimated incidence of medically-attended LRTI cases, LRTI hospital admissions, and excess LRTI-associated deaths preventable by PCV15 and PCV20 under two approaches: (1) comparing PCV15/20 use to no adult PCV immunization, representing the comprehensive effects of implementing these vaccines; and (2) comparing PCV15/20 use to continued PCV13 use under pre-2022 ACIP recommendations, representing the incremental reduction resulting from extended serotype coverage.^11^ We used a vaccine-probe framework to define the proportion of disease preventable by each vaccine, incorporating prior PCV13 direct-effect estimates of 45.6% (95% confidence interval: 21.8-62.5%) for disease associated with PCV13 serotypes,^4^ and 9.5% (2.2-16.3%) for LRTI due to all causes within the study population,^8^ as described in the supporting information (**Text S2**). We used data from a previous US study of LRTI etiology to define the relative frequency of detection of PCV13 and PCV15/20 serotypes within age- and risk-specific strata, and thus estimate the expansion in coverage of LRTI burden expected with the transition from PCV13 to PCV15/20 (**Table S2**). Our analyses assumed no effect of PPSV23 on non-bacteremic LRTI, consistent with prior evidence from the study population.^8^ We estimated total cases preventable within the US population assuming age- and stratum-specific rates resembled those observed among KPSC patients.

## RESULTS

Analyses included 3,681,430 individuals, with 2,893,265 (78.6%) contributing data over ≥2 years (**Table 1**). Throughout follow-up, the proportions of participants contributing person-time in the 18-29, 30-49, 50-64, 65-79, and ≥80 year age brackets at any point were 20.6%, 34.5%, 24.8%, 15.9%, and 4.3%, respectively. Most individuals (69.1%; *n*=2,852,505) belonged to the ACIP low-risk stratum at some point during follow-up. In total, 569,001 individuals aged ≥65 years (84.4% of 674,085) had received PCV13 at any point, while 83.8% (*n*=565,197) and 76.5% (*n*=515,704) had received PPSV23 or both vaccines, respectively. Only 26.6% (*n*=23,313 of 87,762) and 22.3% (*n*=19,547) of high-risk adults aged 18-64 years had received PCV13 and complete a series of PCV13 and PPSV23, respectively, at any point. Among 759,362 at-risk individuals aged 18-64 years, 397,738 (52.4%) had received PPSV23 at any point.

In total, 324,002 (8.8% of 3,681,430) individuals experienced LRTI during follow-up (135,708 [3.7%] CAP and 188,294 [5.1%] non-CAP LRTI; **Table 1**). Compared to persons not experiencing LRTI, cases were older, more likely to belong to the at-risk or high-risk strata, and had higher prevalence of all chronic comorbid conditions assessed and higher rates of healthcare utilization across all care settings. These differences were most pronounced for CAP cases, among whom 22.5% (*n=*29,262 of 135,708) were aged ≥80 years, 45.9% (*n*=62,798) and 21.6% (*n*=28,110) belonged to the at-risk and high-risk strata, respectively, and 17.4% (*n=*23,652) had experienced ≥1 inpatient admission in year preceding their CAP diagnosis.

Incidence rates of any LRTI, non-CAP LRTI, and CAP were 34.1 (29.3-38.3), 20.5 (16.2-26.2), and 14.3 (13.3-15.3), respectively, per 1,000 person-years among all adults aged ≥18 years (**Table 2**). Incidence rates of any LRTI, non-CAP LRTI, and CAP resulting in inpatient admission were 3.7 (3.5-4.0), 0.5 (0.4-0.6), and 3.3 (3.2-3.5) per 1,000 person-years, respectively. Rates for all LRTI spanned 21.7 (15.9-26.7) to 89.9 (83.4-101.8) cases per 1,000 person-years across age groups, and were 23.9 (20.5-27.2), 50.0 (48.7-55.7), and 88.3 (81.9-91.4) cases per 1,000 person-years within the low-risk, at-risk, and high-risk strata, respectively. Among at-risk adults aged 18-64 years, who were not previously recommended to receive PCV13, LRTI incidence exceeded rates among low-risk adults aged 65-79 years (42.5 [38.7-49.8] vs. 27.2 [25.0-30.7] per 1,000 person-years). Incidence rates of hospitalized LRTI and hospitalized CAP were comparable between these groups (2.5 [2.2-3.4] and 2.7 [2.3-3.1] hospitalized LRTI cases per 1,000 person-years, respectively, and 2.1 [1.9-3.0] and 2.5 [2.1-2.9] hospitalized CAP cases per 1,000 person-years, respectively). We present counterfactual rates, estimated under a scenario of no adult immunization with PCV13, in **Table S3**.

**Table 2:**
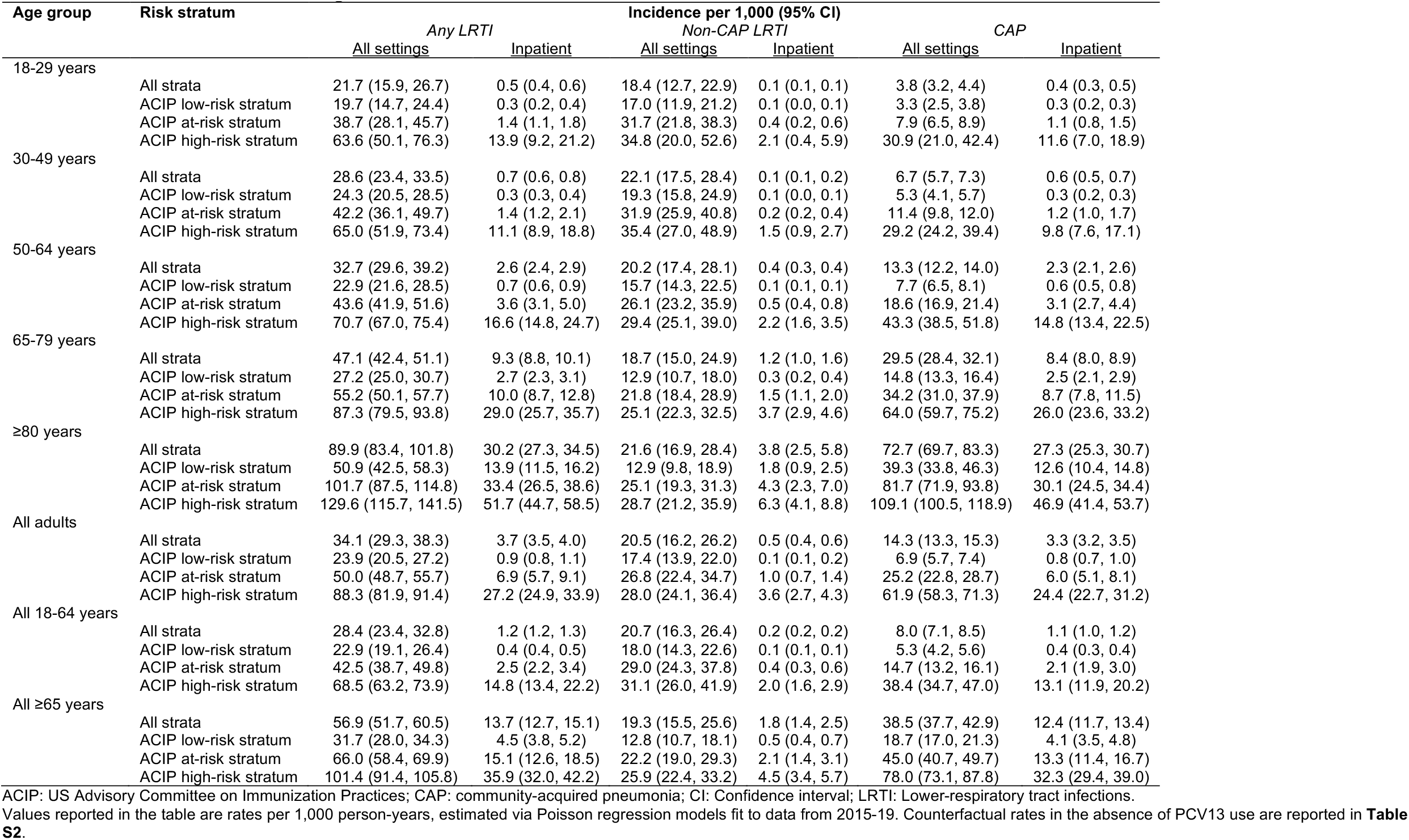
Incidence of LRTI across ages and risk strata.

| Age group | Risk stratum | Incidence per 1,000 (95% CI) |  |  |  |  |  |
| --- | --- | --- | --- | --- | --- | --- | --- |
|  |  | Any LRTI |  | Non-CAP LRTI |  | CAP |  |
|  |  | All settings | Inpatient | All settings | Inpatient | All settings | Inpatient |
| 18-29 years | All strata | 21.7 (15.9, 26.7) | 0.5 (0.4, 0.6) | 18.4 (12.7, 22.9) | 0.1 (0.1, 0.1) | 3.8 (3.2, 4.4) | 0.4 (0.3, 0.5) |
|  | ACIP low-risk stratum | 19.7 (14.7, 24.4) | 0.3 (0.2, 0.4) | 17.0 (11.9, 21.2) | 0.1 (0.0, 0.1) | 3.3 (2.5, 3.8) | 0.3 (0.2, 0.3) |
|  | ACIP at-risk stratum | 38.7 (28.1, 45.7) | 1.4 (1.1, 1.8) | 31.7 (21.8, 38.3) | 0.4 (0.2, 0.6) | 7.9 (6.5, 8.9) | 1.1 (0.8, 1.5) |
|  | ACIP high-risk stratum | 63.6 (50.1, 76.3) | 13.9 (9.2, 21.2) | 34.8 (20.0, 52.6) | 2.1 (0.4, 5.9) | 30.9 (21.0, 42.4) | 11.6 (7.0, 18.9) |
| 30-49 years | All strata | 28.6 (23.4, 33.5) | 0.7 (0.6, 0.8) | 22.1 (17.5, 28.4) | 0.1 (0.1, 0.2) | 6.7 (5.7, 7.3) | 0.6 (0.5, 0.7) |
|  | ACIP low-risk stratum | 24.3 (20.5, 28.5) | 0.3 (0.3, 0.4) | 19.3 (15.8, 24.9) | 0.1 (0.0, 0.1) | 5.3 (4.1, 5.7) | 0.3 (0.2, 0.3) |
|  | ACIP at-risk stratum | 42.2 (36.1, 49.7) | 1.4 (1.2, 2.1) | 31.9 (25.9, 40.8) | 0.2 (0.2, 0.4) | 11.4 (9.8, 12.0) | 1.2 (1.0, 1.7) |
|  | ACIP high-risk stratum | 65.0 (51.9, 73.4) | 11.1 (8.9, 18.8) | 35.4 (27.0, 48.9) | 1.5 (0.9, 2.7) | 29.2 (24.2, 39.4) | 9.8 (7.6, 17.1) |
| 50-64 years | All strata | 32.7 (29.6, 39.2) | 2.6 (2.4, 2.9) | 20.2 (17.4, 28.1) | 0.4 (0.3, 0.4) | 13.3 (12.2, 14.0) | 2.3 (2.1, 2.6) |
|  | ACIP low-risk stratum | 22.9 (21.6, 28.5) | 0.7 (0.6, 0.9) | 15.7 (14.3, 22.5) | 0.1 (0.1, 0.1) | 7.7 (6.5, 8.1) | 0.6 (0.5, 0.8) |
|  | ACIP at-risk stratum | 43.6 (41.9, 51.6) | 3.6 (3.1, 5.0) | 26.1 (23.2, 35.9) | 0.5 (0.4, 0.8) | 18.6 (16.9, 21.4) | 3.1 (2.7, 4.4) |
|  | ACIP high-risk stratum | 70.7 (67.0, 75.4) | 16.6 (14.8, 24.7) | 29.4 (25.1, 39.0) | 2.2 (1.6, 3.5) | 43.3 (38.5, 51.8) | 14.8 (13.4, 22.5) |
| 65-79 years | All strata | 47.1 (42.4, 51.1) | 9.3 (8.8, 10.1) | 18.7 (15.0, 24.9) | 1.2 (1.0, 1.6) | 29.5 (28.4, 32.1) | 8.4 (8.0, 8.9) |
|  | ACIP low-risk stratum | 27.2 (25.0, 30.7) | 2.7 (2.3, 3.1) | 12.9 (10.7, 18.0) | 0.3 (0.2, 0.4) | 14.8 (13.3, 16.4) | 2.5 (2.1, 2.9) |
|  | ACIP at-risk stratum | 55.2 (50.1, 57.7) | 10.0 (8.7, 12.8) | 21.8 (18.4, 28.9) | 1.5 (1.1, 2.0) | 34.2 (31.0, 37.9) | 8.7 (7.8, 11.5) |
|  | ACIP high-risk stratum | 87.3 (79.5, 93.8) | 29.0 (25.7, 35.7) | 25.1 (22.3, 32.5) | 3.7 (2.9, 4.6) | 64.0 (59.7, 75.2) | 26.0 (23.6, 33.2) |
| ≥80 years | All strata | 89.9 (83.4, 101.8) | 30.2 (27.3, 34.5) | 21.6 (16.9, 28.4) | 3.8 (2.5, 5.8) | 72.7 (69.7, 83.3) | 27.3 (25.3, 30.7) |
|  | ACIP low-risk stratum | 50.9 (42.5, 58.3) | 13.9 (11.5, 16.2) | 12.9 (9.8, 18.9) | 1.8 (0.9, 2.5) | 39.3 (33.8, 46.3) | 12.6 (10.4, 14.8) |
|  | ACIP at-risk stratum | 101.7 (87.5, 114.8) | 33.4 (26.5, 38.6) | 25.1 (19.3, 31.3) | 4.3 (2.3, 7.0) | 81.7 (71.9, 93.8) | 30.1 (24.5, 34.4) |
|  | ACIP high-risk stratum | 129.6 (115.7, 141.5) | 51.7 (44.7, 58.5) | 28.7 (21.2, 35.9) | 6.3 (4.1, 8.8) | 109.1 (100.5, 118.9) | 46.9 (41.4, 53.7) |
| All adults | All strata | 34.1 (29.3, 38.3) | 3.7 (3.5, 4.0) | 20.5 (16.2, 26.2) | 0.5 (0.4, 0.6) | 14.3 (13.3, 15.3) | 3.3 (3.2, 3.5) |
|  | ACIP low-risk stratum | 23.9 (20.5, 27.2) | 0.9 (0.8, 1.1) | 17.4 (13.9, 22.0) | 0.1 (0.1, 0.2) | 6.9 (5.7, 7.4) | 0.8 (0.7, 1.0) |
|  | ACIP at-risk stratum | 50.0 (48.7, 55.7) | 6.9 (5.7, 9.1) | 26.8 (22.4, 34.7) | 1.0 (0.7, 1.4) | 25.2 (22.8, 28.7) | 6.0 (5.1, 8.1) |
|  | ACIP high-risk stratum | 88.3 (81.9, 91.4) | 27.2 (24.9, 33.9) | 28.0 (24.1, 36.4) | 3.6 (2.7, 4.3) | 61.9 (58.3, 71.3) | 24.4 (22.7, 31.2) |
| All 18-64 years | All strata | 28.4 (23.4, 32.8) | 1.2 (1.2, 1.3) | 20.7 (16.3, 26.4) | 0.2 (0.2, 0.2) | 8.0 (7.1, 8.5) | 1.1 (1.0, 1.2) |
|  | ACIP low-risk stratum | 22.9 (19.1, 26.4) | 0.4 (0.4, 0.5) | 18.0 (14.3, 22.6) | 0.1 (0.1, 0.1) | 5.3 (4.2, 5.6) | 0.4 (0.3, 0.4) |
|  | ACIP at-risk stratum | 42.5 (38.7, 49.8) | 2.5 (2.2, 3.4) | 29.0 (24.3, 37.8) | 0.4 (0.3, 0.6) | 14.7 (13.2, 16.1) | 2.1 (1.9, 3.0) |
|  | ACIP high-risk stratum | 68.5 (63.2, 73.9) | 14.8 (13.4, 22.2) | 31.1 (26.0, 41.9) | 2.0 (1.6, 2.9) | 38.4 (34.7, 47.0) | 13.1 (11.9, 20.2) |
| All ≥65 years | All strata | 56.9 (51.7, 60.5) | 13.7 (12.7, 15.1) | 19.3 (15.5, 25.6) | 1.8 (1.4, 2.5) | 38.5 (37.7, 42.9) | 12.4 (11.7, 13.4) |
|  | ACIP low-risk stratum | 31.7 (28.0, 34.3) | 4.5 (3.8, 5.2) | 12.8 (10.7, 18.1) | 0.5 (0.4, 0.7) | 18.7 (17.0, 21.3) | 4.1 (3.5, 4.8) |
|  | ACIP at-risk stratum | 66.0 (58.4, 69.9) | 15.1 (12.6, 18.5) | 22.2 (19.0, 29.3) | 2.1 (1.4, 3.1) | 45.0 (40.7, 49.7) | 13.3 (11.4, 16.7) |
|  | ACIP high-risk stratum | 101.4 (91.4, 105.8) | 35.9 (32.0, 42.2) | 25.9 (22.4, 33.2) | 4.5 (3.4, 5.7) | 78.0 (73.1, 87.8) | 32.3 (29.4, 39.0) |

ACIP: US Advisory Committee on Immunization Practices; CAP: community-acquired pneumonia; CI: Confidence interval; LRTI: Lower-respiratory tract infections.

The 30-day, 60-day, 90-day, and 180-day risks of death after any LRTI diagnosis were 13.7, 18.0, 20.6, and 25.7 per 1,000 cases, respectively, across all age and risk strata (**Table 3**). Of this observed mortality, excess deaths associated with LRTI diagnosis spanned 13.1 (13.0-13.2) to 22.2 (22.1-22.3) per 1,000 cases over 30 to 180 days. Excess mortality associated with CAP diagnoses over these intervals spanned 23.3 (23.2-23.4) to 38.7 (35.8-38.9) per 1,000 cases, greatly exceeding excess mortality associated with non-CAP LRTI (**Table S4**; **Table S5**). Excess LRTI-associated mortality was higher in older age groups compared to younger age groups, and in at-risk or high-risk strata compared to low-risk strata. However, in contrast to patterns in disease incidence, observed and excess LRTI-associated mortality among at-risk adults aged 18-64 years was well below that in adults aged 65-79 or ≥80 years within any risk stratum (6.0 [5.8-6.2] to 10.5 [10.3-10.8] per 1,000 LRTI cases among at-risk adults aged 18-64 years; 25.6 [25.0-26.1] to 43.8 [43.1-44.5] per 1,000 LRTI cases among low risk adults aged 65-79 years; **Table 3**).

**Table 3:** Observed and excess risk of death associated with LRTI over 180 days after diagnosis.

| Age group | Risk stratum | Mortality per 1000 cases (95% CI) |  |  |  |  |  |  |  |
| --- | --- | --- | --- | --- | --- | --- | --- | --- | --- |
|  |  | 30 days |  | 60 days |  | 90 days |  | 180 days |  |
|  |  | <u>Observed mortality</u> | <u>Excess mortality</u> | <u>Observed mortality</u> | <u>Excess mortality</u> | <u>Observed mortality</u> | <u>Excess mortality</u> | <u>Observed mortality</u> | <u>Excess mortality</u> |
| 18-29 years | All strata | 1.0 | 0.9 (0.9, 1.0) | 1.3 | 1.2 (1.1, 1.3) | 1.3 | 1.3 (1.2, 1.3) | 1.6 | 1.5 (1.4, 1.5) |
|  | ACIP low-risk stratum | 0.9 | 0.8 (0.8, 0.9) | 1.1 | 1.1 (1.0, 1.1) | 1.1 | 1.1 (1.0, 1.1) | 1.3 | 1.2 (1.1, 1.3) |
|  | ACIP at-risk stratum | 1.5 | 1.4 (1.1, 1.6) | 2.0 | 1.9 (1.6, 2.1) | 2.1 | 1.9 (1.6, 2.2) | 2.5 | 2.1 (1.8, 2.5) |
|  | ACIP high-risk stratum | 9.1 | 8.5 (5.9, 11.1) | 14.1 | 12.7 (9.4, 16.0) | 19.3 | 17.3 (13.4, 21.1) | 26.4 | 22.9 (18.4, 27.5) |
| 30-49 years | All strata | 2.4 | 2.3 (2.2, 2.4) | 2.9 | 2.8 (2.7, 2.9) | 3.3 | 3.1 (3.0, 3.2) | 3.8 | 3.5 (3.4, 3.6) |
|  | ACIP low-risk stratum | 1.8 | 1.7 (1.7, 1.8) | 2.1 | 2.1 (2.0, 2.2) | 2.4 | 2.3 (2.3, 2.4) | 2.7 | 2.6 (2.5, 2.7) |
|  | ACIP at-risk stratum | 2.7 | 2.6 (2.4, 2.8) | 3.6 | 3.5 (3.2, 3.7) | 4.0 | 3.8 (3.5, 4.0) | 4.9 | 4.4 (4.1, 4.6) |
|  | ACIP high-risk stratum | 23.7 | 22.6 (20.8, 24.3) | 28.9 | 26.9 (24.9, 28.9) | 31.1 | 27.9 (25.8, 30.0) | 37.9 | 31.3 (28.9, 33.6) |
| 50-64 years | All strata | 9.8 | 9.5 (9.4, 9.7) | 12.8 | 12.2 (12.0, 12.4) | 14.5 | 13.6 (13.4, 13.8) | 17.5 | 15.8 (15.6, 16.1) |
|  | ACIP low-risk stratum | 7.7 | 7.6 (7.4, 7.8) | 9.7 | 9.5 (9.3, 9.8) | 10.7 | 10.5 (10.2, 10.7) | 12.3 | 11.7 (11.5, 12.0) |
|  | ACIP at-risk stratum | 10.0 | 9.6 (9.3, 9.9) | 13.4 | 12.7 (12.4, 13.1) | 15.2 | 14.2 (13.8, 14.6) | 19.0 | 17.0 (16.6, 17.5) |
|  | ACIP high-risk stratum | 36.2 | 33.6 (32.1, 35.0) | 47.3 | 42.5 (40.8, 44.1) | 55.7 | 48.5 (46.6, 50.3) | 69.2 | 56.0 (53.9, 58.1) |
| 65-79 years | All strata | 32.7 | 31.4 (31.0, 31.8) | 43.0 | 40.5 (40.0, 40.9) | 49.5 | 45.8 (45.3, 46.2) | 62.7 | 55.1 (54.6, 55.7) |
|  | ACIP low-risk stratum | 26.0 | 25.6 (25.0, 26.1) | 33.4 | 32.5 (31.9, 33.1) | 38.1 | 36.8 (36.1, 37.4) | 46.6 | 43.8 (43.1, 44.5) |
|  | ACIP at-risk stratum | 31.9 | 30.7 (30.1, 31.3) | 41.4 | 39.1 (38.4, 39.8) | 47.4 | 43.9 (43.2, 44.6) | 59.5 | 52.3 (51.5, 53.1) |
|  | ACIP high-risk stratum | 61.7 | 57.3 (55.8, 58.8) | 85.3 | 76.5 (74.8, 78.3) | 99.4 | 86.5 (84.6, 88.4) | 131.1 | 106.4 (104.1, 108.6) |
| ≥80 years | All strata | 104.3 | 96.8 (95.5, 98.1) | 138.3 | 123.9 (122.4, 125.4) | 159.7 | 138.5 (136.9, 140.1) | 202.6 | 162.0 (160.1, 163.8) |
|  | ACIP low-risk stratum | 102.6 | 98.8 (96.5, 101.1) | 130.8 | 123.1 (120.5, 125.7) | 146.4 | 134.6 (131.8, 137.3) | 181.6 | 158.5 (155.4, 161.5) |
|  | ACIP at-risk stratum | 96.0 | 88.4 (86.5, 90.2) | 129.7 | 115.3 (113.2, 117.4) | 151.2 | 130.3 (128.0, 132.6) | 193.2 | 152.9 (150.3, 155.5) |
|  | ACIP high-risk stratum | 123.0 | 110.5 (107.4, 113.6) | 165.5 | 141.3 (137.6, 144.8) | 194.2 | 158.6 (154.7, 162.5) | 249.9 | 183.1 (178.7, 187.6) |
| All adults | All strata | 13.7 | 13.1 (13.0, 13.2) | 18.0 | 16.8 (16.7, 16.9) | 20.6 | 18.8 (18.7, 18.9) | 25.7 | 22.2 (22.1, 22.3) |
|  | ACIP low-risk stratum | 7.5 | 7.4 (7.3, 7.5) | 9.5 | 9.2 (9.1, 9.3) | 10.7 | 10.2 (10.1, 10.3) | 12.7 | 11.8 (11.7, 11.9) |
|  | ACIP at-risk stratum | 20.8 | 19.7 (19.5, 20.0) | 27.6 | 25.6 (25.3, 25.8) | 31.7 | 28.7 (28.4, 29.0) | 40.0 | 34.1 (33.7, 34.5) |
|  | ACIP high-risk stratum | 62.5 | 57.6 (56.6, 58.5) | 84.4 | 74.7 (73.5, 75.8) | 98.5 | 84.2 (83.0, 85.4) | 127.2 | 100.1 (98.7, 101.5) |
| All 18-64 years | All strata | 4.4 | 4.2 (4.2, 4.3) | 5.6 | 5.4 (5.3, 5.5) | 6.3 | 6.0 (5.9, 6.1) | 7.6 | 6.9 (6.8, 7.0) |
|  | ACIP low-risk stratum | 3.0 | 2.9 (2.9, 3.0) | 3.7 | 3.6 (3.6, 3.7) | 4.1 | 4.0 (3.9, 4.1) | 4.7 | 4.5 (4.4, 4.5) |
|  | ACIP at-risk stratum | 6.2 | 6.0 (5.8, 6.2) | 8.4 | 7.9 (7.7, 8.1) | 9.4 | 8.8 (8.6, 9.0) | 11.7 | 10.5 (10.3, 10.8) |
|  | ACIP high-risk stratum | 31.4 | 29.3 (28.2, 30.4) | 40.6 | 36.8 (35.5, 38.0) | 47.3 | 41.5 (40.2, 42.9) | 58.7 | 47.9 (46.4, 49.5) |
| All ≥65 years | All strata | 49 | 46.4 (45.9, 46.8) | 64.7 | 59.6 (59.1, 60.1) | 74.6 | 67.1 (66.5, 67.6) | 94.4 | 79.7 (79.1, 80.3) |
|  | ACIP low-risk stratum | 39.3 | 38.3 (37.7, 38.9) | 50.3 | 48.3 (47.6, 49.0) | 57 | 53.9 (53.2, 54.7) | 69.9 | 63.8 (62.9, 64.6) |
|  | ACIP at-risk stratum | 46.9 | 44.2 (43.6, 44.9) | 62.1 | 57.1 (56.4, 57.8) | 71.7 | 64.3 (63.6, 65.1) | 90.7 | 76.1 (75.2, 77.0) |
|  | ACIP high-risk stratum | 82.7 | 75.8 (74.3, 77.2) | 112.6 | 99.0 (97.4, 100.7) | 131.6 | 111.6 (109.8, 113.4) | 171.5 | 133.7 (131.5, 135.8) |

ACIP: US Advisory Committee on Immunization Practices; CI: Confidence interval.

Compared to a scenario of no adult immunization with PCVs, we estimated that PCV15 could prevent 89.3 (41.3-131.8) medically-attended LRTI cases, 21.9 (10.1-32.0) hospital admissions, and 7.1 (3.3-10.5) deaths per 10,000 person-years among adults aged ≥65 years (**Table 4**; **Table S6**). We estimated that PCV20 could prevent 108.6 (50.4-159.1) medically-attended LRTI cases, 26.6 (12.4-38.7) hospital admissions, and 8.7 (4.0-12.7) deaths per 10,000 person-years within the same age groups. The numbers of adults aged ≥65 years needing to be vaccinated (NNV) to prevent ≥1 medically-attended LRTI case over a 5-year timespan were 22 (15-48) for PCV15 and 18 (13-39) for PCV20 (**Table 5**). Switching from PCV13 to PCV15 was expected to prevent 23.6 (11.1-35.8) LRTI cases, 5.6 (2.6-9.0) LRTI hospital admissions, and 2.0 (1.0-3.0) excess LRTI-associated deaths per 10,000 person-years, while switching to PCV20 was expected to prevent 42.4 (20.1-62.1) LRTI cases, 10.4 (4.9-16.1) LRTI hospital admissions, and 3.7 (1.8-5.4) excess LRTI-associated deaths per 10,000 person-years (**Table 6**).

**Table 4:** Burden of LRTI potentially preventable by PCV15 and PCV20, when implemented in differing risk strata.

| Age group | Risk stratum | Incidence prevented per 10,000 (95% CI) |  |  |  |  |  |
| --- | --- | --- | --- | --- | --- | --- | --- |
|  |  | PCV15 (including PCV13 serotypes) |  |  | PCV20 (including PCV13 serotypes) |  |  |
|  |  | Cases | Admissions | Deaths | Cases | Admissions | Deaths |
| 18-29 years | All strata | 42.4 (18.8, 71.3) | 1.0 (0.4, 1.4) | 0.1 (0.0, 0.1) | 50.3 (22.3, 84.3) | 1.1 (0.5, 1.7) | 0.1 (0.0, 0.1) |
|  | ACIP low-risk stratum | 38.0 (16.8, 63.8) | 0.6 (0.3, 1.0) | 0.0 (0.0, 0.1) | 45.5 (20.2, 76.1) | 0.7 (0.3, 1.1) | 0.1 (0.0, 0.1) |
|  | ACIP at-risk stratum | 73.2 (32.6, 121.6) | 2.8 (1.3, 4.5) | 0.2 (0.1, 0.3) | 87.7 (39.1, 144.8) | 3.4 (1.5, 5.3) | 0.2 (0.1, 0.3) |
|  | ACIP high-risk stratum | 132.8 (60.9, 207.0) | 29.0 (12.6, 52.8) | 3.0 (1.3, 5.0) | 154.9 (70.9, 239.8) | 33.8 (14.7, 61.1) | 3.5 (1.6, 5.7) |
| 30-49 years | All strata | 56.7 (25.7, 91.8) | 1.5 (0.7, 2.2) | 0.2 (0.1, 0.3) | 67.3 (30.6, 108.4) | 1.7 (0.8, 2.6) | 0.2 (0.1, 0.4) |
|  | ACIP low-risk stratum | 47.7 (21.8, 76.4) | 0.7 (0.3, 1.0) | 0.1 (0.1, 0.2) | 57.1 (26.2, 91.1) | 0.8 (0.4, 1.2) | 0.1 (0.1, 0.2) |
|  | ACIP at-risk stratum | 83.7 (38.4, 132.0) | 2.9 (1.3, 5.0) | 0.4 (0.2, 0.6) | 100.2 (46.2, 157.2) | 3.5 (1.6, 6.0) | 0.4 (0.2, 0.7) |
|  | ACIP high-risk stratum | 133.6 (61.0, 206.2) | 24.4 (10.9, 47.7) | 4.2 (1.9, 6.5) | 155.5 (71.2, 238.9) | 28.4 (12.8, 55.3) | 4.9 (2.2, 7.5) |
| 50-64 years | All strata | 67.7 (31.3, 103.9) | 5.3 (2.5, 7.8) | 1.1 (0.5, 1.6) | 80.3 (37.4, 122.7) | 6.3 (2.9, 9.1) | 1.3 (0.6, 1.9) |
|  | ACIP low-risk stratum | 47.3 (21.8, 73.8) | 1.5 (0.7, 2.3) | 0.6 (0.3, 0.9) | 56.6 (26.3, 87.8) | 1.8 (0.8, 2.8) | 0.7 (0.3, 1.0) |
|  | ACIP at-risk stratum | 89.5 (41.6, 135.1) | 7.3 (3.4, 12.4) | 1.5 (0.7, 2.3) | 107.4 (50.1, 160.8) | 8.8 (4.0, 14.8) | 1.8 (0.9, 2.7) |
|  | ACIP high-risk stratum | 150.1 (69.6, 221.3) | 36.8 (16.8, 65.1) | 8.4 (3.9, 12.4) | 175.0 (81.9, 256.2) | 42.9 (19.7, 75.6) | 9.8 (4.6, 14.4) |
| 65-79 years | All strata | 74.3 (34.3, 109.8) | 14.9 (6.9, 21.6) | 4.1 (1.9, 6.1) | 90.4 (41.9, 132.7) | 18.1 (8.4, 26.1) | 5.0 (2.3, 7.3) |
|  | ACIP low-risk stratum | 47.0 (21.6, 70.5) | 4.6 (2.1, 7.0) | 2.1 (0.9, 3.1) | 55.6 (25.7, 82.9) | 5.4 (2.5, 8.2) | 2.4 (1.1, 3.6) |
|  | ACIP at-risk stratum | 93.3 (43.2, 137.2) | 17.4 (7.9, 28.2) | 4.9 (2.3, 7.2) | 110.5 (51.4, 161.4) | 20.6 (9.4, 33.2) | 5.8 (2.7, 8.4) |
|  | ACIP high-risk stratum | 122.2 (56.1, 183.1) | 41.6 (18.8, 65.3) | 13.0 (6.0, 19.5) | 154.8 (72.1, 228.7) | 52.6 (24.2, 82.0) | 16.5 (7.7, 24.4) |
| ≥80 years | All strata | 144.2 (66.6, 214.2) | 48.2 (22.3, 71.7) | 23.4 (10.8, 34.7) | 175.1 (81.3, 258.2) | 58.6 (27.1, 86.5) | 28.4 (13.2, 41.8) |
|  | ACIP low-risk stratum | 86.2 (39.5, 131.5) | 23.5 (10.7, 36.3) | 13.7 (6.3, 20.9) | 102.0 (46.9, 154.7) | 27.8 (12.7, 42.7) | 16.2 (7.4, 24.5) |
|  | ACIP at-risk stratum | 172.7 (79.4, 262.0) | 55.6 (25.2, 88.0) | 26.4 (12.1, 40.1) | 204.6 (94.1, 307.7) | 65.8 (29.9, 103.6) | 31.3 (14.4, 47.1) |
|  | ACIP high-risk stratum | 181.5 (83.1, 273.7) | 72.2 (33.0, 110.4) | 33.2 (15.2, 50.2) | 230.2 (106.9, 342.1) | 91.6 (42.1, 138.2) | 42.1 (19.6, 62.8) |
| All adults | All strata | 67.0 (30.8, 100.9) | 7.4 (3.4, 10.7) | 1.5 (0.7, 2.2) | 80.4 (37.1, 120.6) | 8.9 (4.1, 12.8) | 1.8 (0.8, 2.7) |
|  | ACIP low-risk stratum | 43.9 (20.2, 67.4) | 1.7 (0.8, 2.6) | 0.5 (0.2, 0.8) | 52.3 (24.1, 80.0) | 2.0 (0.9, 3.1) | 0.6 (0.3, 0.9) |
|  | ACIP at-risk stratum | 95.3 (44.1, 138.2) | 12.9 (5.9, 21.5) | 3.2 (1.5, 4.7) | 113.5 (52.7, 163.7) | 15.3 (7.0, 25.6) | 3.9 (1.8, 5.6) |
|  | ACIP high-risk stratum | 148.1 (68.7, 215.3) | 47.4 (22.0, 74.0) | 14.8 (6.9, 21.5) | 180.4 (84.0, 260.2) | 57.7 (26.8, 89.4) | 18.1 (8.4, 26.0) |
| All 18-64 years | All strata | 56.5 (25.8, 89.6) | 2.5 (1.2, 3.7) | 0.4 (0.2, 0.6) | 67.1 (30.7, 105.7) | 3.0 (1.4, 4.3) | 0.5 (0.2, 0.7) |
|  | ACIP low-risk stratum | 44.8 (20.4, 70.8) | 0.8 (0.4, 1.3) | 0.2 (0.1, 0.3) | 53.6 (24.6, 84.4) | 1.0 (0.5, 1.5) | 0.2 (0.1, 0.4) |
|  | ACIP at-risk stratum | 85.7 (39.6, 131.5) | 5.1 (2.4, 8.6) | 0.9 (0.4, 1.4) | 102.7 (47.8, 156.7) | 6.2 (2.8, 10.2) | 1.1 (0.5, 1.7) |
|  | ACIP high-risk stratum | 144.7 (67.2, 214.8) | 32.9 (15.1, 58.8) | 6.9 (3.2, 10.3) | 168.8 (78.8, 248.1) | 38.4 (17.7, 68.2) | 8.1 (3.8, 11.9) |
| All ≥65 years | All strata | 89.3 (41.3, 131.8) | 21.9 (10.1, 32.0) | 7.1 (3.3, 10.5) | 108.6 (50.4, 159.1) | 26.6 (12.4, 38.7) | 8.7 (4.0, 12.7) |
|  | ACIP low-risk stratum | 53.6 (24.6, 80.4) | 7.7 (3.5, 11.9) | 3.4 (1.6, 5.1) | 63.5 (29.3, 94.5) | 9.1 (4.2, 14.0) | 4.0 (1.9, 6.0) |
|  | ACIP at-risk stratum | 111.4 (51.3, 164.7) | 25.7 (11.6, 41.6) | 8.5 (3.9, 12.5) | 131.9 (60.8, 193.6) | 30.5 (13.8, 48.8) | 10.0 (4.6, 14.7) |
|  | ACIP high-risk stratum | 140.8 (64.7, 210.5) | 51.0 (23.3, 78.7) | 18.8 (8.6, 28.1) | 178.5 (83.1, 263.0) | 64.6 (29.9, 98.7) | 23.9 (11.1, 35.2) |

ACIP: US Advisory Committee on Immunization Practices; CAP: community-acquired pneumonia; LRTI: Lower-respiratory tract infections; PCV: pneumococcal conjugate vaccine.

**Table 5:**
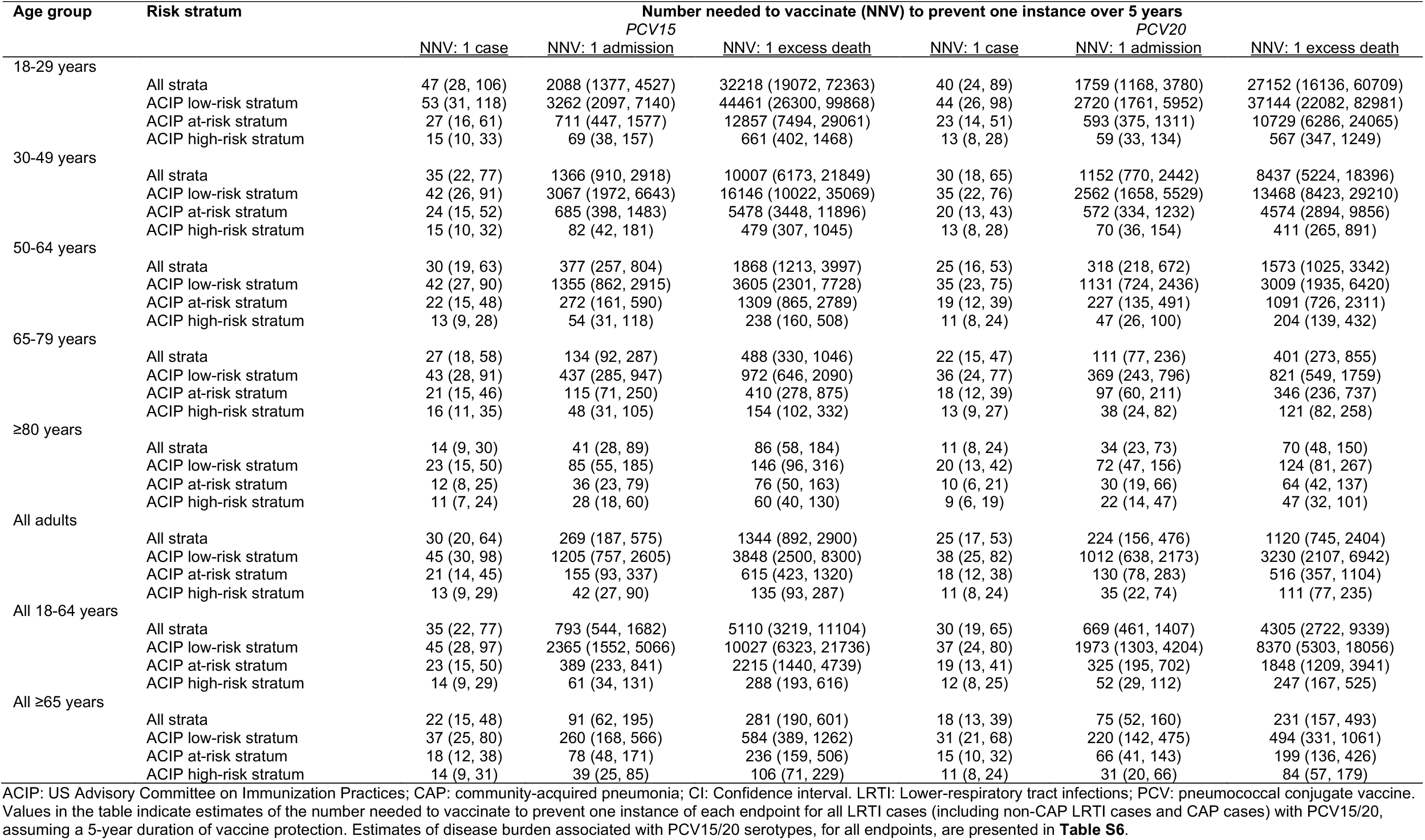
Numbers needed to vaccinate with PCV15 and PCV20 to prevent cases, admissions, and excess deaths.

**Table 6:**
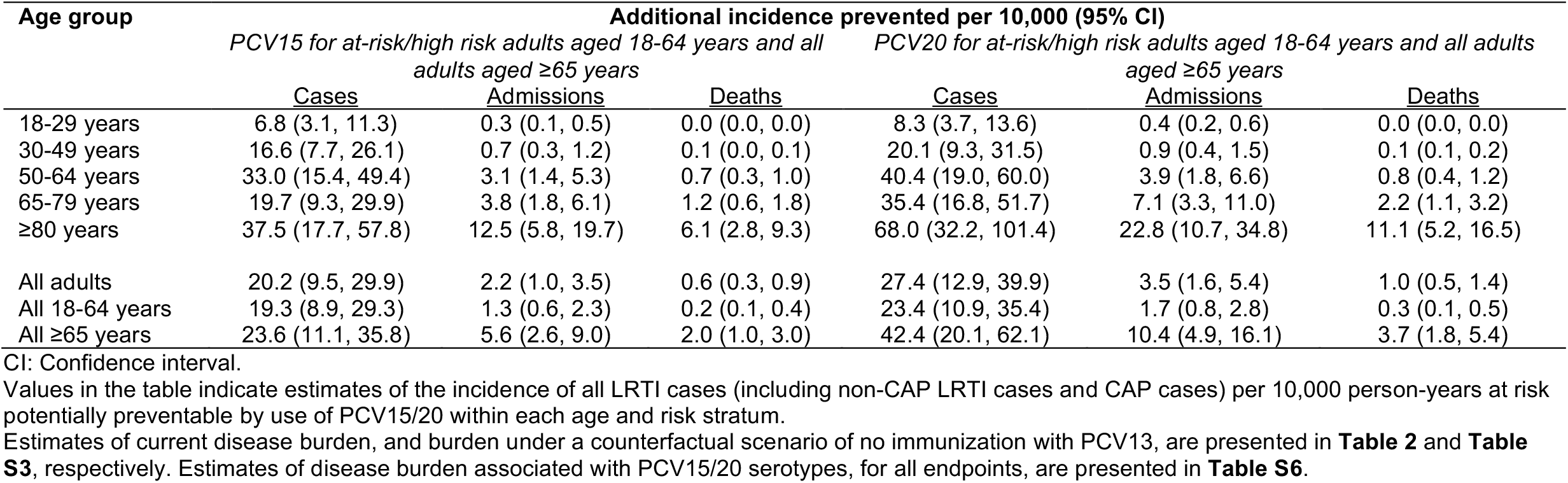
Additional LRTI burden potentially preventable by updated PCV15 and PCV20 recommendations, relative to prior recommendations with PCV13.

Among high-risk adults aged 18-64 years, PCV15 and PCV20, respectively, were expected to prevent 144.7 (67.2-214.8) and 168.8 (78.8-248.1) LRTI cases, 32.9 (15.1-58.8) and 38.4 (17.7-68.2) LRTI hospital admissions, and 6.9 (3.2-10.3) and 8.1 (3.8-11.9) excess LRTI-associated deaths per 10,000 person-years (**Table 4**). For at-risk adults in this age group, who were not previously recommended to receive PCV13, we estimated that PCV15 and PCV20, respectively, could prevent 89.5 (41.6-135.1) and 102.7 (47.8-156.7) LRTI cases, 5.1 (2.4-8.6) and 6.2 (2.8-10.2) LRTI hospital admissions, and 0.9 (0.4-1.4) and 1.1 (0.5-1.7) LRTI-associated excess deaths per 10,000 person-years. Overall, the transition from PCV13 to PCV15 or PCV20 among high-risk adults aged 18-64 years, and new adoption of PCV15 or PCV20 among at-risk adults aged 18-64 years, was expected to prevent an additional 19.3 (8.9-29.3) to 23.4 (10.9-35.4) LRTI cases, 1.3 (0.6-2.3) to 1.7 (0.8-2.8) LRTI hospital admissions, and 0.2 (0.1-0.4) to 0.3 (0.1-0.5) excess LRTI-associated deaths per 10,000 person-years within this age group, relative to pre-existing recommendations with PCV13 (**Table 5**; **Table 6**).

Based on these rates, universal implementation of new recommendations for immunization with PCV15 or PCV20 for all US adults aged ≥65 years, and at-risk or high-risk adults aged <65 years, was expected to more than double the number LRTI cases programmatically preventable by PCVs, relative to continued use of PCV13 under pre-2022 recommendations (up to 967,000 [449,000-1,399,000] cases preventable with PCV15 and 1,159,000 [541,000-1,667,000] cases preventable with PCV20, annually; **Table 7**). This intervention was also expected to increase the number of programmatically-preventable LRTI hospital admissions (166,000 [76,000-258,000] with PCV15 and 200,000 [92,000-309,000] with PCV20, annually) and excess LRTI-associated deaths (58,000 [27,000-84,000] with PCV15 and 70,000 [32,000-101,000] with PCV20, annually). Whereas the expansion of PCV15/20 recommendations to at-risk adults aged <65 years accounted for most of this increase in programmatically-preventable cases, the switch from PCV13 to PCV15/20 among adults aged ≥65 years accounted for most of the increase in programmatically-preventable hospital admissions and deaths.

**Table 7:** LRTI burden targeted by expanded PCV15 and PCV20 recommendations.

| Age group |  | Frequency within US population, per year (95% CI) |  |  |
| --- | --- | --- | --- | --- |
|  |  | Cases | Admissions | Deaths |
|  |  | *n*, in thousands | *n*, in thousands | *n*, in thousands |
| All adults | Estimated total, absent adult PCV immunization | 8,845 (8,117, 9,864) | 1,035 (928, 1,245) | 371 (332, 391) |
|  | Preventable by prior PCV13 recommendations | 437 (199, 645) | 106 (48, 165) | 40 (18, 59) |
|  | Preventable by PCV15 under expanded recommendations | 967 (449, 1,399) | 166 (76, 258) | 58 (27, 84) |
|  | Preventable by PCV20 under expanded recommendations | 1159 (541, 1,667) | 200 (92, 309) | 70 (32, 101) |
| Ages 18-64 years | Estimated total, absent adult PCV immunization | 5,703 (4,963, 6,569) | 245 (229, 330) | 52 (48, 61) |
|  | Preventable by prior PCV13 recommendations | 49 (22, 75) | 11 (5, 20) | 2 (1, 4) |
|  | Preventable by PCV15 under expanded recommendations | 441 (205, 665) | 39 (18, 66) | 8 (3, 11) |
|  | Preventable by PCV20 under expanded recommendations | 526 (246, 790) | 46 (21, 78) | 9 (4, 13) |
| Ages ≥65 years | Estimated total, absent adult PCV immunization | 3,231 (2,895, 3,389) | 797 (688, 921) | 317 (280, 340) |
|  | Preventable by prior PCV13 recommendations | 387 (175, 576) | 94 (43, 147) | 37 (17, 56) |
|  | Preventable by PCV15 under expanded recommendations | 521 (240, 763) | 127 (58, 195) | 50 (23, 74) |
|  | Preventable by PCV20 under expanded recommendations | 626 (291, 911) | 154 (71, 235) | 61 (28, 89) |

CI: Confidence interval.
Values in the table indicate estimates of the all LRTI cases (including non-CAP LRTI cases and CAP cases) preventable within the US population, under the assumption of age-specific incidence rates resembling those observed within the KPSC study population.

## DISCUSSION

New recommendations for use of PCV15/20 may prevent up to 20.2-27.4 additional LRTI cases, 2.2-3.5 additional LRTI hospital admissions, and 0.6-1.0 additional excess LRTI-associated deaths per 10,000 person-years among all US adults aged ≥18 years. Across all age- and risk-based strata for whom PCV15 and PCV20 are now recommended, we estimate that ≥1 medically-attended LRTI case can be prevented for every 18-23 adults who receive PCV15 or PCV20; that ≥1 hospitalized LRTI case can be prevented for every 75-389 adults who are vaccinated; and that ≥1 excess LRTI-associated death can be prevented for every 231-2,215 adults who are vaccinated. Coverage of additional serotypes with PCV15/20 among adults aged ≥65 years accounts for most of the expected increase in hospitalizations and mortality that may be programmatically preventable. However, broadening of PCV15/20 recommendations to at-risk, immunocompetent adults aged <65 years with comorbid conditions—many of whom were not previously eligible for PCV13—accounts for most of the anticipated impact on medically-attended LRTI cases. Whereas prior assessments have emphasized prevention of severe infections including hospitalized CAP and IPD,^18–20^ prevention of outpatient-attended illness may also afford considerable health economic value.^21,22^

Our findings add to growing evidence of the substantial burden of disease preventable by direct immunization of adults with PCVs. Prior vaccine effectiveness studies in KPSC,^8^ elsewhere within the US,^6,7^ and in other countries^9,10^ with mature pediatric PCV13 programs have estimated that direct immunization with PCV13 prevents 7-12% of all-cause LRTI or CAP among adults. These estimates exceed the proportion of cases attributed to PCV13-serotype pneumococci in etiology studies using either conventional culture^2^ or urinary antigen-based diagnostic approaches,^23,24^ suggesting such methods have limitations.^25^ Sensitivity of urinary antigen detection assays for pneumococcal serotype identification in CAP has been validated in cases with culture-confirmed bacteremic pneumonia,^24^ but remains unknown for non-bacteremic cases, underscoring the challenge of defining vaccine-preventable LRTI burden with available diagnostic methodologies.

Prevention of virus-associated LRTI by PCVs has also been reported among both adults^26^ and children.^27^ Facilitative interactions between pneumococci and viruses in the upper or lower respiratory tract may contribute to these observations.^28–32^ Our study has limitations. First, although KPSC members are representative of regional demographic patterns,^16^ members may have better health status and lower risk of LRTI than the US population at large. Among US Medicare beneficiaries aged ≥65 years,^6^ 25.6% of individuals belonged to the ACIP low-risk stratum, in comparison to 49.3% of KPSC members aged ≥65 years in our study; moreover, 39.1% were immunocompromised, in comparison to 17.7% within our sample.

Thus, our study likely underestimates LRTI burden within the broader US population. Second, data on serotype distributions in LRTI came from a sentinel surveillance study outside of KPSC. Differences may exist in the clinical spectrum and characteristics of cases enrolled in this study versus among patients diagnosed under real-world prescribing practices at KPSC. Third, while PCV15 and PCV20 have proven to be safe and immunogenic in adults, real-world effectiveness against LRTI in adults remains to be determined. Correlates of immune protection against mucosal pneumococcal disease endpoints including LRTI are unknown, and the lack of PPSV23-associated protection against non-bacteremic LRTI among adults suggests IgG and opsonophagocytic antibody responses may be imperfect proxies for clinical protection.^8,33^ Whereas minor variation in antibody responses to shared serotypes across PCV7/10/13 has not been associated with differences in clinical protection among children,^34,35^ it remains important to determine whether the same holds true for PCV15/20 among adults. Fourth, residual confounding from unmeasured risk factors or imperfectly-measured risk factors may undermine our estimates of LRTI-associated mortality. However, our analytic framework improves on standard approaches, which may be prone to either over-estimation (e.g., attributing all deaths to LRTI within a certain time window following diagnosis) or under-estimation (e.g., monitoring for in-hospital deaths only) of LRTI mortality.^36^ Last, it remains to be determined how pediatric immunization with PCV15 and PCV20 may impact vaccine-preventable disease burden among adults. The incremental benefit of direct immunization with PCV15/20 among adults may decline if use in children substantially reduces circulation of PCV15/20 serotypes. Nonetheless, considerable burden of PCV13-preventable LRTI has persisted among adults in settings with mature PCV7/13 pediatric immunization programs.^6–8,15,23^

Data from our study complement other assessments of the burden of disease preventable by PCV15 and PCV20, including surveillance studies of serotypes isolated from IPD cases^37^ and serotypes detected via urinary antigen shedding among adults with LRTI.^23^ Our findings support new ACIP recommendations for transitioning from use of PCV13 to PCV15/20 among adults, and implementing PCV15/20 among at-risk adults aged <65 years, as strategies to mitigate LRTI burden. Long-term monitoring of the impact of these recommendations remains crucial to optimize next-generation PCV implementation in the US, and to inform adoption among adults in other settings.

## Data Availability

Anonymized data that support the findings of this study may be made available from the investigative team in the following conditions: (1) agreement to collaborate with the study team on all publications, (2) provision of external funding for administrative and investigator time necessary for this collaboration, (3) demonstration that the external investigative team is qualified and has documented evidence of training for human subjects protections, and (4) agreement to abide by the terms outlined in data use agreements between institutions.

## ACKNOWLEDGMENTS

The authors thank Joann Zamparo for assistance in this work.

## Text S1: Counterfactual framework for estimating excess LRTI-associated risk of death

To estimate counterfactual risks of death for cases, we estimated risk of mortality over 30, 60, 90, and 180-day periods among “control” individuals who did not experience CAP or non-CAP LRTI within the same calendar year as cases, reweighting observations among these individuals according to their propensity for case status based on age group, sex, race/ethnicity, body mass index, cigarette smoking, neighborhood socioeconomic status, ACIP-defined risk group, history of healthcare utilization across outpatient, inpatient, and ED settings in the prior year, history of LRTI in the prior year, vaccines received, and comorbid conditions (as categorized in **Table 1** for all listed variables). We generated propensity weights for case status using logistic models fitted via lasso penalized regression, incorporating all covariates listed above. To account for seasonality in LRTI diagnoses and all-cause mortality, we sampled start dates for 30, 60, 90, and 180-day mortality surveillance periods among controls at random from the distribution of diagnosis dates for cases, each year. We defined excess LRTI-associated mortality (for endpoints of any LRTI, pneumonia, and non-pneumonia LRTI), overall and within each age- and risk-specific stratum, via the risk difference among cases versus controls. We fit risk differences on the absolute scale via linear regression models accounting for case propensity weights.

## Text S2: Vaccine impact estimation

Whereas PCV13 efficacy and effectiveness against vaccine-serotype CAP has been established in earlier randomized^1^ and observational^2^ studies, approval of PCV15 and PCV20 was granted based on non-inferior immunogenicity relative to PCV13 and PPSV23 for overlapping serotypes in prelicensure trials.^3,4^ There is no established correlate of protection against pneumococcal CAP or IPD among adults. However, among trial participants who received PCV15 or PCV20, opsonophagocytic antibody responses for PCV13-targeted serotypes were of similar magnitude to responses for the added serotypes (22F and 33F for PCV15; 8, 10A, 11A, 12F, 15B/C, 22F, and 33F for PCV20). Based on this finding, as well as experience with PCV7, PCV10, and PCV13 in pediatric programs,^5,6^ we assumed protection conferred by PCV15 and PCV20 against LRTI due to serotypes targeted by each vaccine would be equivalent to serotype-specific protection conferred by PCV13.^1^

Defining *λ* as the incidence of disease due to all causes, *p*_13_ as the proportion of cases associated with PCV13-serotype pneumococci (absent vaccination), and *θ* as the risk (or rate) ratio of PCV13-serotype disease resulting from vaccine-conferred protection,^1^ the incidence of disease preventable by direct vaccination with PCV13 was expected to be equal to *θλp*_13_. We used the proportion of disease associated with PCV13 serotypes (*π*_13_), under observed PCV13 coverage (*ν*), to define

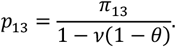

We estimated *π*_13_ in a vaccine-probe framework, accounting for protection against the specific endpoint of PCV13-serotype disease^1^ (*θ*, estimated to be 0.544 [95% confidence interval: 0.375-0.782]) and the risk ratio of all-cause disease associated with PCV13 receipt in the study population^7^ (*ω*, estimated to be 0.905 [0.837-0.978]). As derived elsewhere,^8^ we considered

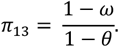

We used the observed proportion of cases associated with PCV15 and PCV20 serotypes (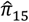 and 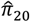), as compared to PCV13 serotypes (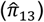), to define the expected incidence of disease preventable by these vaccines, relative to no vaccination, as 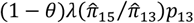 and 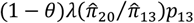, respectively. While accommodating potential under-detection of PCV13-serotype disease via the vaccine probe estimation methodology for *π*_13_, this framework assumed equivalent sensitivity of serotype-specific urinary antigen detection assays for PCV13-targeted and non-PCV13 serotypes, such that

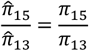

and

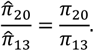

We defined the incidence of disease preventable by PCV15 and PCV20, relative to PCV13, as 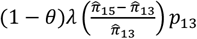 and 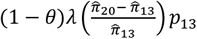, respectively.

To propagate statistical uncertainty, we sampled from fitted distributions of *λ, θ*, and *ω*, as defined in the main-text methods sections and references cited. We defined Dirichlet distributions for 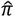. parameterized by the number of cases with PCV13 serotypes, serotypes 22F and 33F, serotypes 8, 10A, 11A, 12F, 15B/C, and no pneumococcal serotypes, observed in each age- and risk-specific stratum.^9^

We also estimated the Number of individuals Needing to be Vaccinated with PCV15/20 (NNV_15_ and NNV_20_) to prevent ≥1 LRTI case, ≥1 hospitalized LRTI case, and ≥1 death over a 5-year timespan. We defined this timeframe based on prior estimates of the minimum duration of PCV13-derived protection against vaccine-serotype CAP^1,7^ and again assuming similar performance characteristics for PCV15/20; true values of the NNV_15_ and NNV_20_ may thus exceed our estimates if at least partial protection extends >5 years after vaccination. We defined the NNV for each vaccine as the inverse of the expected per-capita reduction in LRTI cases, hospital admissions, and attributable deaths over a 5-year span associated with vaccination:

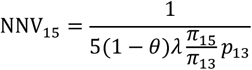

and

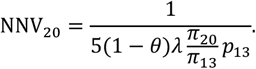

Last, we projected our estimates of age-specific incidence rates of LRTI, hospital admissions, and excess mortality to census estimates of the 2021 US population size (based on interannual projections^10^) to estimate the total number of cases, admissions, and excess deaths preventable by PCV15 and PCV20. As direct estimates of estimates of the number of individuals within each age stratum belonging to ACIP-defined low-risk, at-risk, and high-risk strata are not available, we assumed the distribution of these risk groups across age groups resembled that among KPSC members included in the analysis. As other studies of large administrative databases have revealed higher prevalence of chronic comorbid conditions and immunocompromise, in comparison to that observed within KPSC,^11^ we expected any discrepancy would lead to under-estimation of the true burden of LRTI preventable by PCV15 and PCV20.

---

**Table S1:** ICD-10 codes for outcome assignment.

| ICD-10-CM code | Definition | Outcome |  |
| --- | --- | --- | --- |
|  |  | Non-CAP LRTI | CAP |
| J09– | Influenza due to certain identified influenza viruses | X |  |
| J09.X1 | With pneumonia |  | X |
| J10– | Influenza due to other identified influenza virus | X |  |
| J10.00 | With unspecified type of pneumonia |  | X |
| J10.01 | With the same other identified influenza virus pneumonia |  | X |
| J10.08 | With other unspecified pneumonia |  | X |
| J11– | Influenza due to unidentified influenza virus | X |  |
| J11.00 | With unspecified type of pneumonia |  | X |
| J11.08 | With specified pneumonia |  | X |
| J12– | Viral pneumonia, not elsewhere classified |  | X |
| J13– | Pneumonia due to *Streptococcus pneumoniae* |  | X |
| J14 | Pneumonia due to *Haemophilus influenzae* |  | X |
| J15– | Bacterial pneumonia, not elsewhere classified |  | X |
| J16– | Pneumonia due to other infectious organisms, not elsewhere classified |  | X |
| J17 | Pneumonia in diseases classified elsewhere |  | X |
| J18– (excl. J18.2) | Pneumonia, unspecified organism (excl. hypostatic pneumonia, unspecified organism) |  | X |
| J20– | Acute bronchitis | X |  |
| J21– | Acute bronchiolitis | X |  |
| J22 | Unspecified acute lower respiratory infection | X |  |

CAP: community-acquired pneumonia; LRTI: Lower-respiratory tract infections.
Items marked JX– are nonspecific (non-billable) codes for which the non-pneumonia LRTI and community-acquired pneumonia classifications indicated in the right columns encompass all sub-codes. For J09–, J10–, and J11–, we designate the specific codes used to differentiate pneumonia diagnoses from other LRTI diagnoses.

**Table S2:** Serotype coverage of PCV13, PCV15, and PCV20 across ages and risk strata.

| Serotypes | Age and risk group, n (%) |  |  |  |
| --- | --- | --- | --- | --- |
|  | 18-64 years |  | ≥65 years |  |
|  | At risk/low risk | High risk | At risk/low risk | High risk |
| All pneumococcal cases | N=308 | N=212 | N=401 | N=190 |
| PCV13 serotypes (1, 3, 4, 5, 6A, 6B, 7F, 9V, 14, 18C, 19A, 19F, 23F) | 163 (52.9) | 106 (50.0) | 194 (48.4) | 96 (50.5) |
| Additional PCV15 serotypes (22F, 33F) | 61 (198) | 37 (17.5) | 87 (21.7) | 49 (25.8) |
| Additional PCV20 serotypes (8, 10A, 11A, 12F, 15B) | 43 (14.0) | 40 (18.9) | 58 (14.5) | 25 (13.2) |
| Non-PCV serotypes | 41 (13.3) | 29 (13.7) | 62 (15.5) | 20 (10.5) |

Data on serotype coverage are obtained from Isturiz R, Grant L, Gray S, et al. Expanded analysis of 20 pneumococcal serotypes associated with radiographically confirmed community-acquired pneumonia in hospitalized US adults. *Clin Infect Dis* 2021; 73: 1216–22.

**Table S3:**
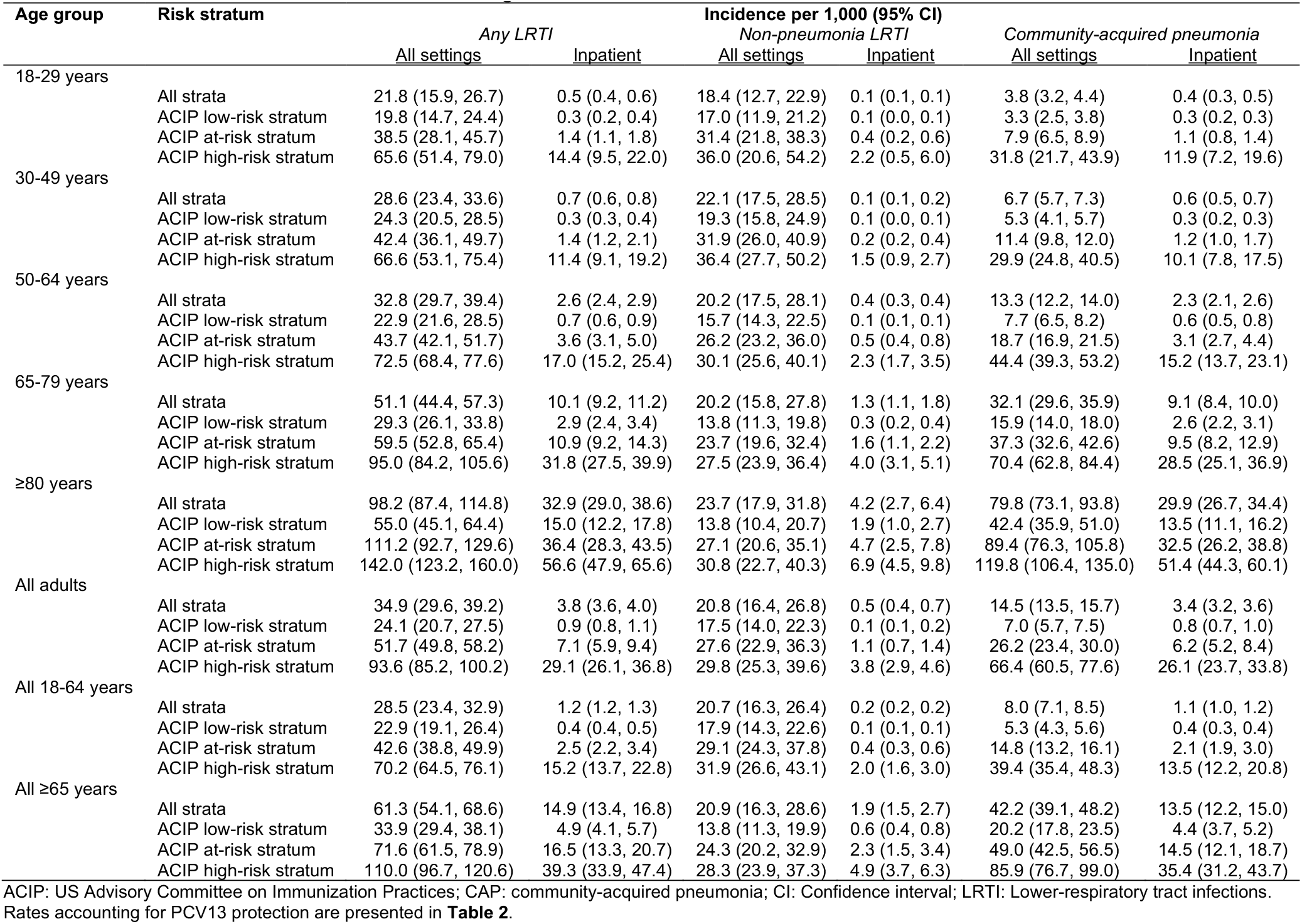
Estimated incidence of LRTI across ages and risk strata, under a scenario of no adult immunization with PCVs.

**Table S4:** Excess mortality associated with LRTI cases not presenting with community-acquired pneumonia (non-CAP LRTI).

| Age group | Risk stratum | Mortality per 1,000 cases (95% CI) |  |  |  |  |  |  |  |
| --- | --- | --- | --- | --- | --- | --- | --- | --- | --- |
|  |  | 30 days |  | 60 days |  | 90 days |  | 180 days |  |
|  |  | <u>Observed mortality</u> | <u>Excess mortality</u> | <u>Observed mortality</u> | <u>Excess mortality</u> | <u>Observed mortality</u> | <u>Excess mortality</u> | <u>Observed mortality</u> | <u>Excess mortality</u> |
| 18-29 years | All strata | 0.1 | -- | 0.1 | -- | 0.1 | -- | 0.2 | -- |
|  | ACIP low-risk stratum | -- | -- | -- | -- | -- | -- | -- | -- |
|  | ACIP at-risk stratum | 0.3 | 0.3 (0.1, 0.4) | 0.9 | 0.7 (0.5, 0.9) | 0.9 | 0.7 (0.5, 0.9) | 1.0 | 0.6 (0.4, 0.9) |
|  | ACIP high-risk stratum | 1.2 | 0.7 (-0.4, 1.9) | 1.2 | 0.1 (-1.3, 1.4) | 3.4 | 1.7 (-0.3, 3.6) | 3.4 | 0.4 (-1.8, 2.5) |
| 30-49 years | All strata | 0.3 | 0.3 (0.2, 0.3) | 0.4 | 0.3 (0.3, 0.4) | 0.5 | 0.3 (0.3, 0.4) | 0.7 | 0.4 (0.4, 0.5) |
|  | ACIP low-risk stratum | 0.3 | 0.2 (0.2, 0.3) | 0.4 | 0.3 (0.3, 0.4) | 0.4 | 0.4 (0.3, 0.4) | 0.6 | 0.5 (0.4, 0.5) |
|  | ACIP at-risk stratum | 0.4 | 0.3 (0.2, 0.4) | 0.6 | 0.4 (0.3, 0.5) | 0.6 | 0.3 (0.2, 0.4) | 0.9 | 0.4 (0.2, 0.5) |
|  | ACIP high-risk stratum | 2.1 | 1.1 (0.4, 1.7) | 2.1 | 0.2 (-0.5, 0.9) | 2.4 | -0.5 (-1.3, 0.3) | 6.9 | 0.8 (-0.5, 2.1) |
| 50-64 years | All strata | 1.4 | 1.1 (1.0, 1.2) | 2.1 | 1.5 (1.4, 1.6) | 2.4 | 1.6 (1.5, 1.7) | 3.6 | 2.0 (1.8, 2.1) |
|  | ACIP low-risk stratum | 1.1 | 1.0 (0.9, 1.1) | 1.5 | 1.3 (1.2, 1.4) | 1.6 | 1.4 (1.3, 1.5) | 2.1 | 1.5 (1.4, 1.6) |
|  | ACIP at-risk stratum | 1.4 | 1.1 (1.0, 1.2) | 1.8 | 1.2 (1.0, 1.3) | 2.3 | 1.4 (1.2, 1.5) | 3.8 | 1.9 (1.7, 2.1) |
|  | ACIP high-risk stratum | 5.4 | 3.1 (2.4, 3.7) | 10.6 | 6.2 (5.3, 7.1) | 11.9 | 5.5 (4.5, 6.5) | 18.9 | 7.0 (5.7, 8.3) |
| 65-79 years | All strata | 5.0 | 3.8 (3.6, 4.0) | 7.9 | 5.6 (5.4, 5.8) | 9.9 | 6.5 (6.2, 6.7) | 15.5 | 8.4 (8.1, 8.8) |
|  | ACIP low-risk stratum | 1.7 | 1.3 (1.1, 1.4) | 3.2 | 2.3 (2.1, 2.5) | 4.0 | 2.7 (2.5, 3.0) | 7.3 | 4.6 (4.2, 4.9) |
|  | ACIP at-risk stratum | 5.1 | 4.0 (3.8, 4.3) | 7.8 | 5.6 (5.3, 5.9) | 9.6 | 6.3 (5.9, 6.7) | 14.8 | 8.0 (7.6, 8.5) |
|  | ACIP high-risk stratum | 14.7 | 10.8 (10.0, 11.6) | 23.4 | 15.6 (14.5, 16.6) | 29.3 | 17.9 (16.6, 19.1) | 43.1 | 20.9 (19.4, 22.4) |
| ≥80 years | All strata | 28.4 | 21.6 (20.8, 22.4) | 42.4 | 29.1 (28.1, 30.1) | 52.5 | 33.1 (31.9, 34.2) | 78.0 | 40.5 (39.1, 41.9) |
|  | ACIP low-risk stratum | 23.2 | 19.5 (18.3, 20.8) | 33.4 | 25.9 (24.4, 27.4) | 40.3 | 28.9 (27.3, 30.6) | 56.7 | 34.5 (32.4, 36.5) |
|  | ACIP at-risk stratum | 25.1 | 18.2 (17.1, 19.3) | 38.7 | 25.5 (24.1, 26.9) | 47.7 | 28.5 (26.9, 30.0) | 74.2 | 36.9 (34.9, 38.9) |
|  | ACIP high-risk stratum | 41.6 | 30.2 (28.1, 32.3) | 61.9 | 39.8 (37.2, 42.4) | 79.3 | 46.9 (43.9, 49.9) | 113.4 | 52.2 (48.5, 55.9) |
| All adults | All strata | 2.6 | 2.1 (2.0, 2.1) | 4.0 | 2.9 (2.8, 3.0) | 4.9 | 3.3 (3.2, 3.3) | 7.3 | 4.1 (4.0, 4.2) |
|  | ACIP low-risk stratum | 1.0 | 0.9 (0.9, 0.9) | 1.5 | 1.2 (1.2, 1.2) | 1.8 | 1.3 (1.3, 1.4) | 2.6 | 1.7 (1.6, 1.7) |
|  | ACIP at-risk stratum | 4.1 | 3.1 (3.0, 3.3) | 6.2 | 4.3 (4.2, 4.5) | 7.6 | 4.8 (4.7, 5.0) | 11.7 | 6.3 (6.0, 6.5) |
|  | ACIP high-risk stratum | 15.7 | 11.4 (10.8, 11.9) | 24.6 | 16.0 (15.3, 16.7) | 30.9 | 18.2 (17.4, 19.0) | 45.2 | 21.1 (20.1, 22.1) |
| All 18-64 years | All strata | 0.6 | 0.5 (0.5, 0.5) | 0.9 | 0.7 (0.6, 0.7) | 1.0 | 0.7 (0.7, 0.7) | 1.5 | 0.8 (0.8, 0.9) |
|  | ACIP low-risk stratum | 0.4 | 0.4 (0.3, 0.4) | 0.5 | 0.5 (0.4, 0.5) | 0.6 | 0.5 (0.5, 0.5) | 0.8 | 0.6 (0.5, 0.6) |
|  | ACIP at-risk stratum | 0.9 | 0.7 (0.6, 0.8) | 1.2 | 0.8 (0.7, 0.9) | 1.5 | 0.9 (0.8, 1.0) | 2.3 | 1.1 (1.0, 1.3) |
|  | ACIP high-risk stratum | 4.3 | 2.5 (2.0, 2.9) | 7.9 | 4.4 (3.7, 5.0) | 9.2 | 4.0 (3.2, 4.7) | 14.7 | 4.9 (4.0, 5.9) |
| All ≥65 years | All strata | 10.3 | 7.9 (7.7, 8.1) | 15.7 | 11.1 (10.8, 11.3) | 19.4 | 12.6 (12.3, 12.9) | 29.3 | 15.9 (15.5, 16.3) |
|  | ACIP low-risk stratum | 5.7 | 4.7 (4.5, 5.0) | 8.5 | 6.6 (6.3, 6.9) | 10.5 | 7.5 (7.2, 7.9) | 15.9 | 10.0 (9.6, 10.4) |
|  | ACIP at-risk stratum | 9.9 | 7.5 (7.2, 7.8) | 15.1 | 10.5 (10.1, 10.9) | 18.5 | 11.7 (11.3, 12.2) | 28.6 | 15.2 (14.6, 15.8) |
|  | ACIP high-risk stratum | 23.3 | 17.2 (16.3, 18.1) | 35.7 | 23.6 (22.5, 24.7) | 45.3 | 27.5 (26.2, 28.7) | 65.4 | 31.4 (29.8, 32.9) |

ACIP: US Advisory Committee on Immunization Practices; CI: Confidence interval.

**Table S5:** Excess mortality associated with community-acquired pneumonia (CAP).

| Age group | Risk stratum | Mortality per 1000 cases (95% CI) |  |  |  |  |  |  |  |
| --- | --- | --- | --- | --- | --- | --- | --- | --- | --- |
|  |  | 30 days |  | 60 days |  | 90 days |  | 180 days |  |
|  |  | <u>Observed mortality</u> | <u>Excess mortality</u> | <u>Observed mortality</u> | <u>Excess mortality</u> | <u>Observed mortality</u> | <u>Excess mortality</u> | <u>Observed mortality</u> | <u>Excess mortality</u> |
| 18-29 years | All strata | 4.2 | 4.2 (4.0, 4.3) | 5.6 | 5.6 (5.4, 5.7) | 5.8 | 5.7 (5.6, 5.9) | 6.6 | 6.5 (6.3, 6.6) |
|  | ACIP low-risk stratum | 4.1 | 4.1 (4.0, 4.2) | 5.7 | 5.7 (5.5, 5.8) | 5.8 | 5.7 (5.6, 5.9) | 6.4 | 6.3 (6.1, 6.5) |
|  | ACIP at-risk stratum | 4.1 | 4.0 (3.6, 4.4) | 4.7 | 4.5 (4.1, 5.0) | 5.0 | 4.7 (4.3, 5.2) | 6.0 | 5.6 (5.1, 6.1) |
|  | ACIP high-risk stratum | 21.2 | 20.2 (16.3, 24.0) | 29.6 | 28.0 (23.4, 32.6) | 40.6 | 38.6 (33.2, 43.9) | 53.2 | 49.7 (43.6, 55.8) |
| 30-49 years | All strata | 7.7 | 7.6 (7.5, 7.7) | 9.3 | 9.2 (9.1, 9.4) | 10.5 | 10.3 (10.1, 10.4) | 11.8 | 11.5 (11.3, 11.7) |
|  | ACIP low-risk stratum | 7.0 | 7.0 (6.8, 7.1) | 8.4 | 8.3 (8.2, 8.5) | 9.4 | 9.3 (9.2, 9.5) | 10.3 | 10.1 (10.0, 10.3) |
|  | ACIP at-risk stratum | 8.2 | 8.1 (7.8, 8.4) | 10.7 | 10.5 (10.2, 10.9) | 12.2 | 11.9 (11.6, 12.3) | 14.4 | 13.8 (13.4, 14.2) |
|  | ACIP high-risk stratum | 44.6 | 43.4 (41.0, 45.8) | 54.5 | 52.1 (49.4, 54.7) | 57.6 | 54.3 (51.5, 57.0) | 69.5 | 63.4 (60.4, 66.4) |
| 50-64 years | All strata | 22.6 | 22.3 (22.1, 22.6) | 28.8 | 28.2 (27.9, 28.5) | 32.1 | 31.2 (30.9, 31.5) | 37.5 | 35.8 (35.5, 36.1) |
|  | ACIP low-risk stratum | 20.8 | 20.7 (20.4, 21.0) | 25.8 | 25.6 (25.3, 26.0) | 28.2 | 27.9 (27.6, 28.3) | 31.6 | 31.0 (30.6, 31.4) |
|  | ACIP at-risk stratum | 22.0 | 21.6 (21.2, 22.1) | 28.8 | 28.1 (27.6, 28.6) | 32.1 | 31.1 (30.6, 31.6) | 38.9 | 37.0 (36.4, 37.6) |
|  | ACIP high-risk stratum | 59.6 | 57.2 (55.4, 59.0) | 76.2 | 71.7 (69.6, 73.7) | 88.8 | 82.4 (80.2, 84.6) | 107.2 | 94.6 (92.1, 97.0) |
| 65-79 years | All strata | 50.1 | 48.8 (48.3, 49.3) | 65.1 | 62.6 (62.0, 63.1) | 74.5 | 70.8 (70.2, 71.3) | 92.7 | 85.2 (84.6, 85.9) |
|  | ACIP low-risk stratum | 43.7 | 43.3 (42.6, 44.0) | 56.1 | 55.1 (54.4, 55.9) | 63.8 | 62.4 (61.6, 63.2) | 76.9 | 74.0 (73.1, 74.9) |
|  | ACIP at-risk stratum | 49.1 | 47.9 (47.2, 48.6) | 63.3 | 60.9 (60.1, 61.7) | 72.0 | 68.6 (67.7, 69.4) | 88.4 | 81.4 (80.4, 82.3) |
|  | ACIP high-risk stratum | 81.9 | 77.4 (75.7, 79.1) | 111.3 | 102.7 (100.8, 104.7) | 128.6 | 116.1 (113.9, 118.2) | 168.7 | 144.7 (142.3, 147.2) |
| ≥80 years | All strata | 125.6 | 118.6 (117.2, 120.0) | 165.1 | 151.5 (149.9, 153.1) | 189.4 | 169.3 (167.6, 171.1) | 237.8 | 198.2 (196.2, 200.1) |
|  | ACIP low-risk stratum | 123.8 | 120.1 (117.6, 122.6) | 157.1 | 149.7 (146.9, 152.4) | 175.0 | 163.8 (160.9, 166.7) | 216.3 | 193.7 (190.5, 197.0) |
|  | ACIP at-risk stratum | 116.7 | 109.8 (107.8, 111.8) | 156.2 | 142.7 (140.4, 144.9) | 180.7 | 160.6 (158.1, 163.1) | 227.7 | 188.3 (185.5, 191.1) |
|  | ACIP high-risk stratum | 144.6 | 132.4 (129.1, 135.7) | 192.1 | 169.2 (165.4, 172.9) | 224.1 | 191.0 (186.9, 195.1) | 285.3 | 220.4 (215.7, 225.0) |
| All adults | All strata | 23.9 | 23.3 (23.2, 23.4) | 30.8 | 29.6 (29.5, 29.8) | 34.8 | 33.1 (32.9, 33.2) | 42.1 | 38.7 (38.5, 38.9) |
|  | ACIP low-risk stratum | 15.8 | 15.7 (15.5, 15.8) | 19.9 | 19.6 (19.5, 19.8) | 22.0 | 21.6 (21.4, 21.7) | 25.5 | 24.6 (24.5, 24.8) |
|  | ACIP at-risk stratum | 33.6 | 32.6 (32.2, 32.9) | 44.0 | 42.0 (41.7, 42.4) | 50.1 | 47.2 (46.8, 47.5) | 61.6 | 55.8 (55.3, 56.2) |
|  | ACIP high-risk stratum | 87.1 | 82.1 (81.0, 83.2) | 115.6 | 106.2 (104.9, 107.5) | 133.8 | 120.3 (118.9, 121.7) | 170.6 | 144.4 (142.8, 146.0) |
| All 18-64 years | All strata | 11.6 | 11.5 (11.4, 11.6) | 14.7 | 14.4 (14.3, 14.5) | 16.2 | 15.9 (15.8, 16.0) | 18.7 | 18.0 (17.9, 18.2) |
|  | ACIP low-risk stratum | 9.7 | 9.7 (9.6, 9.8) | 12.1 | 12.0 (11.9, 12.1) | 13.1 | 13.0 (12.9, 13.2) | 14.6 | 14.4 (14.2, 14.5) |
|  | ACIP at-risk stratum | 14.7 | 14.5 (14.2, 14.8) | 19.3 | 18.8 (18.5, 19.1) | 21.6 | 20.9 (20.6, 21.2) | 26 | 24.8 (24.4, 25.1) |
|  | ACIP high-risk stratum | 54.3 | 52.3 (50.9, 53.7) | 69.0 | 65.2 (63.6, 66.7) | 79.2 | 73.9 (72.1, 75.5) | 95.9 | 85.6 (83.7, 87.5) |
| All ≥65 years | All strata | 68.2 | 65.7 (65.2, 66.2) | 89.2 | 84.2 (83.7, 84.8) | 102.1 | 94.8 (94.2, 95.4) | 127.5 | 113.2 (112.5, 113.8) |
|  | ACIP low-risk stratum | 57.9 | 56.0 (55.3, 56.7) | 72.9 | 70.9 (70.1, 71.7) | 82.4 | 79.4 (78.6, 80.3) | 100.2 | 94.1 (93.1, 95.0) |
|  | ACIP at-risk stratum | 65.6 | 63.1 (62.4, 63.8) | 85.9 | 81.1 (80.2, 81.9) | 98.5 | 91.3 (90.4, 92.1) | 122.2 | 107.9 (106.9, 108.9) |
|  | ACIP high-risk stratum | 104.3 | 97.5 (95.9, 99.0) | 140.3 | 127.2 (125.4, 129.0) | 162.7 | 143.8 (141.8, 145.8) | 210.5 | 173.8 (171.6, 176.0) |

ACIP: US Advisory Committee on Immunization Practices; CI: Confidence interval.

**Table S6:** Estimated burden of LRTI associated with PCV15 and PCV20 serotypes.

| Age group | Risk stratum | Incidence associated with indicated serotypes, per 10,000 |  |  |  |  |  |
| --- | --- | --- | --- | --- | --- | --- | --- |
|  |  | PCV15 serotypes |  |  | PCV20 serotypes |  |  |
|  |  | Cases | Admissions | Excess deaths | Cases | Admissions | Excess deaths |
| 18-29 years | All strata | 96.7 (66.4, 129.4) | 2.1 (1.7, 2.6) | 0.1 (0.1, 0.2) | 114.6 (79.1, 152.7) | 2.5 (2.0, 3.1) | 0.2 (0.1, 0.2) |
|  | ACIP low-risk stratum | 86.5 (59.5, 117.2) | 1.4 (1.0, 1.7) | 0.1 (0.1, 0.1) | 103.5 (71.6, 139.6) | 1.6 (1.2, 2.1) | 0.1 (0.1, 0.2) |
|  | ACIP at-risk stratum | 167.2 (115.8, 220.1) | 6.3 (4.5, 8.4) | 0.4 (0.2, 0.5) | 200.4 (139.4, 261.4) | 7.5 (5.4, 10.0) | 0.4 (0.3, 0.6) |
|  | ACIP high-risk stratum | 296.6 (221.0, 383.4) | 65.2 (41.1, 103.3) | 6.8 (4.7, 9.4) | 345.6 (258.6, 444.2) | 76.0 (48.3, 119.7) | 7.9 (5.5, 10.8) |
| 30-49 years | All strata | 125.9 (96.4, 164.0) | 3.2 (2.6, 3.9) | 0.4 (0.3, 0.6) | 149.1 (114.9, 193.5) | 3.8 (3.1, 4.7) | 0.5 (0.4, 0.7) |
|  | ACIP low-risk stratum | 106.0 (81.4, 138.7) | 1.5 (1.1, 1.9) | 0.3 (0.2, 0.4) | 126.7 (98.0, 164.7) | 1.7 (1.3, 2.2) | 0.3 (0.3, 0.4) |
|  | ACIP at-risk stratum | 185.7 (144.5, 239.6) | 6.3 (5.0, 9.5) | 0.8 (0.6, 1.1) | 221.8 (174.1, 285.0) | 7.6 (6.0, 11.4) | 1.0 (0.8, 1.3) |
|  | ACIP high-risk stratum | 299.3 (222.8, 377.9) | 52.8 (38.8, 92.2) | 9.3 (6.9, 12.0) | 349.1 (261.7, 437.4) | 61.5 (45.5, 106.8) | 10.9 (8.1, 13.8) |
| 50-64 years | All strata | 148.1 (122.3, 190.4) | 11.7 (9.9, 13.9) | 2.3 (1.9, 3.0) | 175.6 (145.9, 224.8) | 13.9 (11.8, 16.3) | 2.8 (2.3, 3.6) |
|  | ACIP low-risk stratum | 102.8 (85.6, 136.7) | 3.2 (2.6, 4.3) | 1.2 (1.0, 1.6) | 123.0 (103.1, 162.7) | 3.9 (3.1, 5.1) | 1.4 (1.2, 1.9) |
|  | ACIP at-risk stratum | 196.4 (164.3, 247.3) | 15.9 (12.6, 23.2) | 3.3 (2.8, 4.2) | 235.1 (198.1, 294.4) | 19.0 (15.2, 27.7) | 4.0 (3.4, 5.0) |
|  | ACIP high-risk stratum | 332.4 (272.2, 399.1) | 79.6 (62.5, 124.2) | 18.6 (15.2, 22.5) | 387.5 (320.2, 460.3) | 92.6 (73.4, 143.7) | 21.7 (17.9, 25.9) |
| 65-79 years | All strata | 164.7 (135.4, 195.5) | 32.8 (27.8, 38.2) | 9.1 (7.5, 10.8) | 200.2 (165.8, 236.0) | 39.9 (34.1, 46.1) | 11.0 (9.1, 13.0) |
|  | ACIP low-risk stratum | 103.9 (84.1, 127.9) | 10.1 (7.9, 12.7) | 4.5 (3.7, 5.6) | 122.8 (100.4, 150.3) | 12.0 (9.5, 15.0) | 5.4 (4.4, 6.6) |
|  | ACIP at-risk stratum | 206.7 (170.1, 245.2) | 38.1 (29.8, 52.3) | 10.8 (8.9, 12.8) | 244.7 (202.9, 288.3) | 45.1 (35.5, 61.6) | 12.8 (10.6, 15.1) |
|  | ACIP high-risk stratum | 270.8 (217.8, 329.9) | 91.7 (71.6, 121.5) | 28.8 (23.1, 35.1) | 343.2 (279.7, 411.6) | 116.1 (92.0, 152.5) | 36.5 (29.8, 43.8) |
| ≥80 years | All strata | 317.4 (263.5, 385.7) | 106.4 (87.9, 129.4) | 51.4 (42.6, 62.5) | 385.6 (322.0, 465.8) | 129.3 (107.6, 156.3) | 62.5 (52.2, 75.5) |
|  | ACIP low-risk stratum | 192.1 (148.6, 239.4) | 52.2 (40.3, 66.4) | 30.4 (23.5, 38.0) | 227.5 (177.1, 282.0) | 61.8 (48.1, 78.0) | 36.1 (28.0, 44.7) |
|  | ACIP at-risk stratum | 384.2 (302.0, 475.0) | 124.6 (92.4, 160.4) | 58.8 (46.2, 72.6) | 455.0 (359.8, 558.1) | 147.7 (110.1, 188.5) | 69.6 (55.0, 85.4) |
|  | ACIP high-risk stratum | 402.7 (320.2, 494.9) | 160.4 (125.3, 201.8) | 73.8 (58.5, 90.8) | 510.2 (411.6, 618.1) | 203.4 (160.9, 252.1) | 93.4 (75.3, 113.4) |
| All adults | All strata | 149.2 (119.2, 180.1) | 16.4 (14.1, 18.7) | 3.3 (2.6, 4.0) | 179.2 (143.4, 215.3) | 19.7 (17.0, 22.3) | 4.0 (3.2, 4.8) |
|  | ACIP low-risk stratum | 97.4 (77.6, 120.7) | 3.6 (2.9, 4.8) | 1.2 (0.9, 1.4) | 116.0 (92.9, 143.0) | 4.3 (3.5, 5.7) | 1.4 (1.1, 1.7) |
|  | ACIP at-risk stratum | 209.5 (179.6, 246.1) | 28.1 (22.0, 39.6) | 7.1 (6.1, 8.4) | 249.4 (215.3, 291.9) | 33.5 (26.3, 47.0) | 8.5 (7.3, 10.0) |
|  | ACIP high-risk stratum | 328.5 (276.8, 380.6) | 103.7 (84.9, 136.7) | 32.9 (27.7, 38.1) | 399.4 (338.7, 458.9) | 125.9 (103.9, 165.2) | 40.0 (33.9, 46.0) |
| All 18-64 years | All strata | 126.2 (96.7, 160.4) | 5.6 (4.7, 6.5) | 0.9 (0.7, 1.1) | 149.6 (115.2, 189.4) | 6.6 (5.6, 7.6) | 1.0 (0.8, 1.3) |
|  | ACIP low-risk stratum | 100.0 (76.5, 128.6) | 1.9 (1.5, 2.4) | 0.4 (0.3, 0.6) | 119.6 (92.1, 152.7) | 2.2 (1.8, 2.8) | 0.5 (0.4, 0.7) |
|  | ACIP at-risk stratum | 188.8 (153.1, 239.9) | 11.1 (9.0, 16.0) | 2.0 (1.6, 2.5) | 225.9 (184.1, 285.3) | 13.3 (10.8, 19.1) | 2.4 (1.9, 3.0) |
|  | ACIP high-risk stratum | 320.8 (260.8, 388.2) | 71.0 (56.0, 112.0) | 15.4 (12.5, 18.7) | 374.0 (306.7, 448.5) | 82.5 (66.1, 129.7) | 17.9 (14.7, 21.6) |
| All ≥65 years | All strata | 197.6 (163.6, 234.4) | 48.3 (40.5, 57.1) | 15.8 (13.0, 18.7) | 240.1 (200.3, 282.5) | 58.6 (49.5, 68.9) | 19.1 (16.0, 22.5) |
|  | ACIP low-risk stratum | 118.9 (95.6, 145.1) | 17.0 (13.3, 21.6) | 7.6 (6.1, 9.2) | 140.6 (113.9, 170.3) | 20.2 (15.8, 25.5) | 9.0 (7.3, 10.9) |
|  | ACIP at-risk stratum | 247.2 (199.9, 294.8) | 57.2 (43.3, 76.2) | 18.8 (15.2, 22.4) | 292.8 (238.2, 346.5) | 67.6 (51.6, 89.6) | 22.3 (18.1, 26.4) |
|  | ACIP high-risk stratum | 312.2 (250.5, 378.4) | 112.8 (88.3, 145.0) | 41.7 (33.5, 50.6) | 395.5 (321.9, 472.0) | 142.9 (113.5, 181.4) | 52.9 (43.1, 63.1) |

ACIP: US Advisory Committee on Immunization Practices; CAP: community-acquired pneumonia; LRTI: Lower-respiratory tract infections; PCV: pneumococcal conjugate vaccine.

